# RetFit: A Novel Deep Learning-Based Biomarker of Cardiorespiratory Fitness Derived From the Retina

**DOI:** 10.1101/2025.09.29.25336874

**Authors:** Ian Quintas, Dennis Bontempi, Sacha Bors, Olga Trofimova, Leah Böttger, Ilaria Iuliani, Sofía Ortín Vela, Bart Liefers, Jose Vargas-Quiros, Caroline C.W. Klaver, Ilenia Meloni, Adham Elwakil, Ciara Bergin, Mattia Tomasoni, VascX Consortium, Sven Bergmann, David M. Presby

## Abstract

Cardiorespiratory fitness (CRF) is a strong predictor of cardiovascular events and all-cause mortality, often outperforming traditional risk factors. However, its clinical assessment remains limited due to the need for specialized equipment, personnel, and time demands. Because CRF is closely tied to vascular health, surrogate measures that capture vascular features may provide a practical alternative for its estimation. Retinal Color Fundus Images (CFIs) provide a non-invasive window into systemic vascular health and have already proven useful in predicting cardiovascular risk factors and diseases. However, CFIs have yet to be explored for their potential to predict CRF. In this study, we introduce RetFit, a novel CRF estimator derived from CFIs by leveraging state-of-the-art vision transformers. We evaluated RetFit’s clinical relevance by analyzing its associations with cardiovascular risk factors and disease outcomes, and exploring its genetic architecture, benchmarking it against a submaximal-exercise-test CRF (SETCRF) estimate. RetFit was prognostic of both cardiovascular events (hazard ratios as low as 0.668, 95%CI 0.617–0.723, p<0.001) and overall mortality (hazard ratios as low as 0.780, 95%CI 0.754–0.801, p<0.001), and significantly associated with the majority of disease states and risk factors explored, with these effects being consistent across two external and independent cohorts. Although RetFit and SETCRF shared a moderate phenotypic correlation (*r*=0.45), their significant genetic associations were disjoint. Interpretability analyses suggest a role for retinal vasculature in RetFit’s predictions, with attention maps emphasizing vascular regions and segmentation analyses showing arterial bifurcation count as the strongest associated feature (β=0.287, 95% CI 0.263–0.311, p<0.001). These findings highlight the potential of retinal imaging as a scalable, cost-effective, and accessible alternative for CRF estimation, supporting its use in large-scale screening and risk stratification in both clinical and public health contexts.

## Introduction

Cardiorespiratory fitness (CRF) is a well-established, powerful, and independent predictor of cardiovascular events, mortality, and a wide range of disease outcomes^1,2^. Notably, CRF’s predictive capacity often surpasses that of traditional risk factors such as smoking, hypertension, hypercholesterolemia, and type 2 diabetes, establishing CRF as one of the most powerful markers for clinical risk stratification^1,3–6^. Importantly, CRF is modifiable through lifestyle interventions, such as increased physical activity, and improvements in CRF are associated with reduced risks of cardiovascular events and overall mortality^7,8^. Therefore, efficiently and accurately estimating CRF can play a crucial role in the assessment of overall health status and disease risk. However, current gold-standard methods for measuring CRF are resource-intensive, requiring specialized equipment and trained personnel, which limits their feasibility in routine clinical and population-level settings.

One of the most widely used and informative measures of CRF is maximal oxygen uptake (VO₂max), which is defined as the highest rate at which an individual can consume oxygen during intense exercise. The gold standard for measuring VO₂max is the maximal cardiopulmonary exercise test (CPET)^1,9,10^, which provides an objective and precise assessment of aerobic capacity by continuously measuring respiratory gas exchange during exercise to volitional exhaustion. However, despite its precision, CPET remains underutilized due to significant practical and clinical limitations: it requires expensive equipment, trained personnel, and a maximal volitional effort from the subject, and is contraindicated in subjects with certain cardiovascular predispositions or known conditions (e.g., active endocarditis, decompensated heart failure, unstable angina, physical/mental disability impairing movement)^10–12^. Consequently, submaximal- and non-exercise methods for estimating VO₂max have been developed and have demonstrated diagnostic and prognostic value for various health outcomes^13^. Submaximal exercise tests estimate CRF by modelling the relationship between heart rate (HR) response and work rate^14^, whereas non-exercise estimates of CRF may be derived from phenotypic, demographic, and anthropometric variables, providing a rapid and cost-effective alternative suited for public health and clinical settings^13,15^. In particular, estimates that utilize cardiovascular metrics, like resting heart rate, demonstrate improved performance over estimates without cardiovascular measures, potentially due to the cardiorespiratory system having a central, rate-limiting role in determining VO_2_max^16,17^. Therefore, identifying novel cardiovascular proxies may lead to improvements in the accuracy and accessibility of CRF estimates.

Color fundus images (CFIs) represent a potentially promising source for such novel cardiovascular proxies, as CFIs have been shown to provide valuable insights into the cardiovascular system and reflect systemic vascular health^18,19^. Studies have shown that CFIs, whether analyzed directly via end-to-end deep learning pipelines or through extracted features quantifying retinal vascular properties, can reliably predict multiple cardiovascular risk factors (e.g., age, sex, smoking status, and hypertension)^20,21^ and diseases (e.g., myocardial infarction, ischaemic stroke, and heart failure)^22,23^. Importantly, CFIs are noninvasive and relatively easy to acquire, making them an appealing resource to estimate CRF. Building on this rationale, we develop RetFit, a novel estimator of CRF from retinal images, based on a state-of-the-art Vision Transformer (**Fig. 1a**). To determine the clinical relevance of RetFit, we assessed its associations with genetics, risk factors, and diseases, and directly benchmarked the diagnostic and prognostic capacities of this novel biomarker against an established measure of submaximal exercise testing CRF (SETCRF)^24^ (**Fig. 1b**). Furthermore, we assessed RetFit’s robustness and generalizability by externally validating the biomarker in two external and independent cohorts^25,26^ in which CRF was not measured (**Fig. 1c**). By showing consistent results in the risk factor and disease association analyses, and strong prognostic power through the survival analyses, we prove that RetFit captures clinically meaningful information even in the absence of fitness-based measurements. Ultimately, this work aims to provide a scalable and accessible method for estimating CRF, overcoming the key barriers of conventional testing.

**Figure 1.**
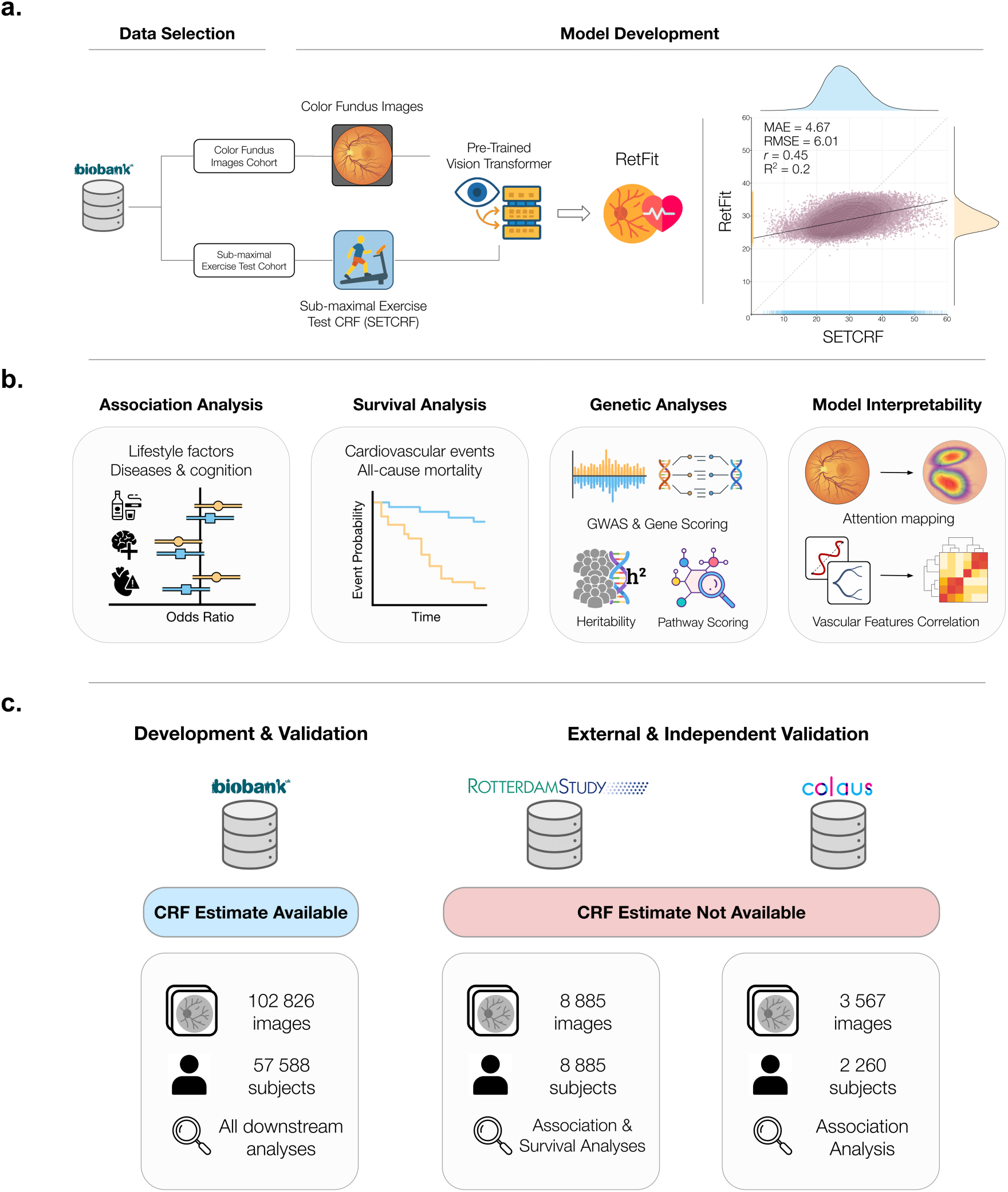
Study Overview. **a)** We selected the subset of genotyped subjects from the UK Biobank who underwent a cardiorespiratory fitness (CRF) assessment via sub-maximal exercise testing (SETCRF) and had colour fundus images (CFIs) taken, as well as access to available health outcome information and risk factors, for a total of N=57 588 subjects and 102 826 images. We fine-tuned a state-of-the-art Vision Transformer to predict CRF from CFIs and obtained a CFI-based estimation of CRF, namely RetFit (16 outlier points not shown on scatter plot for displaying purposes). **b)** Using SETCRF as a benchmark in the UK Biobank dataset, we studied the association of RetFit with common lifestyle factors and diseases and assessed its prognostic power using survival analysis. Furthermore, we investigated the genetic architecture of both RetFit and SETCRF using GWAS, gene enrichment analyses, pathway scoring, and heritability estimation. We examined associations between retinal vascular properties and RetFit/SETCRF using attention mapping. **c)** Finally, we externally validated RetFit in two independent cohorts for which no CRF estimate was collected, the Rotterdam Study (N=8 885, 8 885 images; The Netherlands) and the CoLaus study (N= 2 260, 3 567 images; Switzerland), by running association and survival analyses.

## Results

### Generation of a Fitness Marker from Fundus Images

We fine-tuned a transformer model specifically designed for CFIs, RETFound^22^, on 102 826 images taken from 57 588 participants in the UK Biobank (UKBB) who had performed a sub-maximal exercise test to estimate their CRF (SETCRF). The CFI-derived estimates of CRF, termed RetFit, demonstrated a moderate, positive correlation with SETCRF (*r*=0.45, p<0.001; R^2^=0.20; MAE=4.67; RMSE=6.01; **Fig. 1a**). We then conducted additional analyses to evaluate RetFit’s diagnostic and prognostic value, examine its genetic architecture, and benchmark these associations against SETCRF (**Fig. 1b**). The fine-tuned weights of the transformer model were then used to predict RetFit in two different cohorts, the Rotterdam Study (RS) and the CoLaus Study (CoLaus) (**Fig. 1c**).

### RetFit Informs on Risk Factors and Diseases

We performed a series of regression analyses to evaluate the relative and independent contributions of RetFit and SETCRF to diseases and associated risk factors in the UKBB. First, we assessed their independent associations by building separate models with either RetFit or SETCRF and extracting their standardized coefficients. All the models were adjusted for sex, age, body mass index (BMI), and socioeconomic status to account for confounding effects. We then used log-likelihood ratio tests (LRT) to compare models that included both RetFit and SETCRF to those that included just one or the other to determine whether each predictor offered a unique, non-redundant contribution. To assess the generalizability of RetFit, we ran similar linear regression analyses on the two external and independent cohorts.

Of the 21 binary health outcomes examined, RetFit significantly informed upon 20 of them, whereas SETCRF significantly informed upon 15 outcomes (**Fig. 2a**). RetFit and SETCRF informed upon all binary health outcomes in the same direction except for the current smoking status (against non-smokers) and dyslipidemia. Furthermore, we found five associations were significant for RetFit but not for SETCRF (cataract, cataract surgery, current smoking status, atherosclerosis, and prospective memory). Macular degeneration was the only outcome for which neither of the biomarkers was significantly associated. The LRT results for the binary outcomes showed a significant improvement in model fit when adding both RetFit and SETCRF for cataract, cataract surgery, cancer, atherosclerosis, and prospective memory. As for the logistic regression models, macular degeneration was the only disease for which adding either of the two biomarkers did not result in a significant gain of information. Cancer diagnosis was the only outcome for which adding SETCRF to a model with RetFit improved the model, whereas for all the other outcomes, adding RetFit improved the model with SETCRF (**Fig. 2b**).

**Figure 2.**
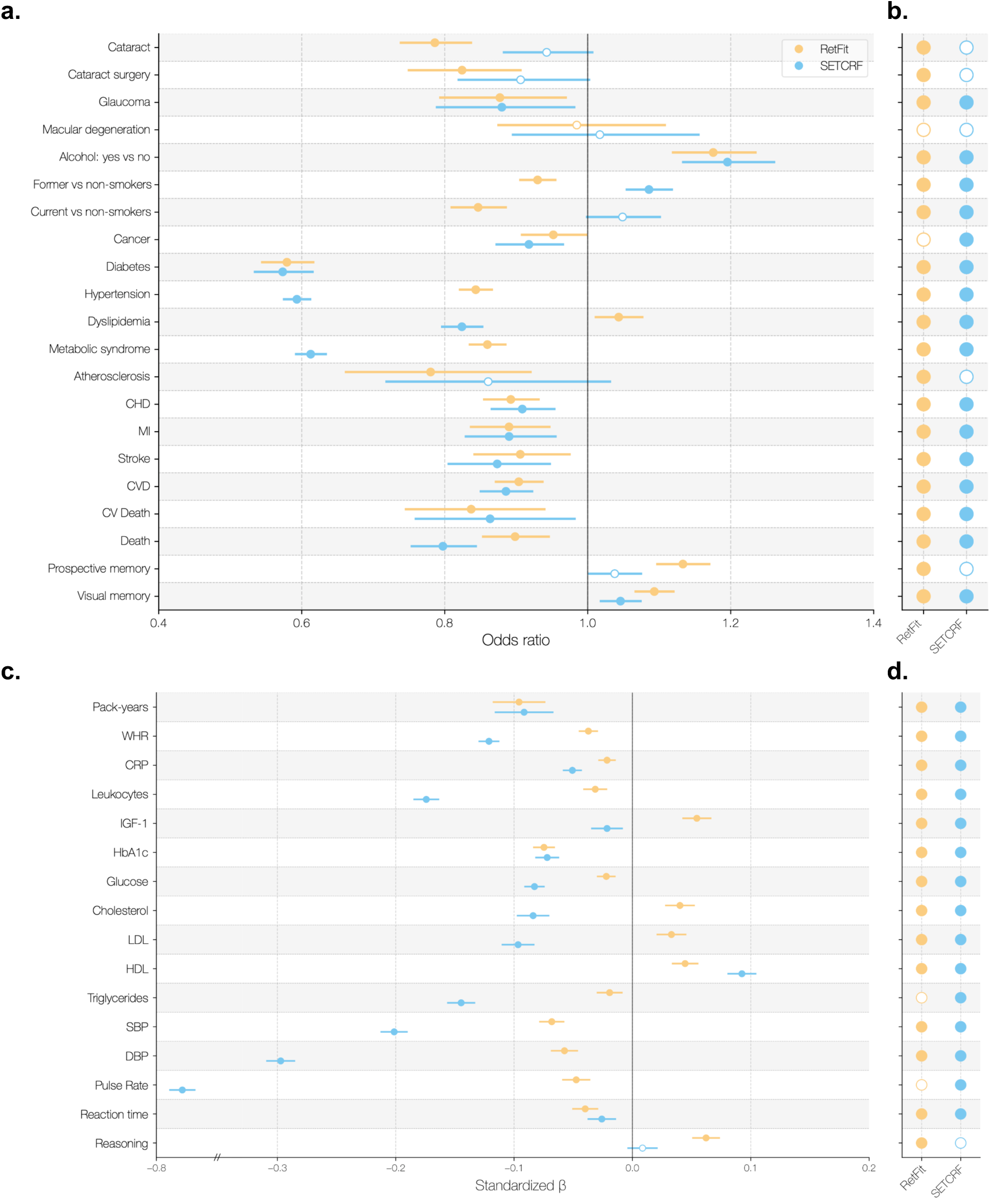
Association of RetFit and SETCRF with diseases and risk factors in the UKBB dataset. **a)** The left panel shows standardized coefficients for RetFit and SETCRF from independent models predicting binary outcomes in the UKBB. **b)** The right panel shows results from likelihood ratio tests (LRTs), comparing models that included both RetFit and SETCRF, with models including only one of the two CRF estimates. **c)** The left panel shows standardized coefficients for RetFit and SETCRF from independent models predicting continuous outcomes. **d)** The right panel shows results from likelihood ratio tests (LRTs), comparing models that included both RetFit and SETCRF with models excluding either CRF estimate. Filled circles indicate significance after Benjamini-Hochberg correction (in both panels). CHD: Coronary Heart Disease; MI: Myocardial Infarction; CVD: Cardiovascular Disease; CV Death: Cardiovascular Death; WHR: waist-to-hip ratio; CRP: C-reactive protein; IGF-1: Insulin-like growth factor 1; HbA1c: Glycated hemoglobin; LDL & HDL: low- and high-density lipoprotein; SBP & DBP: systolic and diastolic blood pressure.

Of the 15 continuous risk factors examined in the UKBB, we found that RetFit was significantly associated with all of them, while SETCRF showed significant associations with all but reasoning (**Fig. 2c**). Of the 14 factors significantly associated with both RetFit and SETCRF, 10 demonstrated concordant directionality. Only insulin-like growth factor 1 (IGF-1), low-density lipoprotein cholesterol (LDL), and total cholesterol exhibited opposing associations, being positively associated with RetFit and negatively associated with SETCRF. These results are consistent with results obtained for dyslipidemia in the binary setting, as we also found opposing directions. Results from the log-likelihood ratio tests demonstrate that reasoning and triglycerides were the only two outcomes for which adding one predictor to a model already containing the other led to a significant improvement in model fit (**Fig. 2d**).

To further validate RetFit’s ability to inform upon disease and investigate its performance in populations without CRF testing available, we performed regression analyses using RetFit estimates derived from the CFIs in the Rotterdam Study (RS) and the CoLaus study (N=8 885 and N = 2 260, respectively) using outcomes analogous to those in the initial analysis shown in **Figure 2a & 2c** (**Fig. 3a & 3b**). Adjusting for sex, age, and BMI, we found that, for the binary disease outcomes, the external validation confirmed most of the significant results obtained in the UKBB in either the RS or in the CoLaus (10/12 and 7/15 significant results reproduced, respectively), with cataract disease and former smokers failing to reach significance in the external validation cohorts (**Fig. 3a**). Moreover, we identified a novel association in the RS between RetFit and macular degeneration (OR_macular deg._=0.853, 95% CI 0.783 to 0.929, p<0.001) that did not reach statistical significance in the UKBB. Regarding the continuous risk factors (**Fig. 3b**), the effect sizes from the external validation reproduced those we obtained in the UKBB in at least one of the two external and independent cohorts (8/10 and 4/10 significant results reproduced for the RS and the CoLaus respectively), except for leukocytes, whose association with RetFit was trending towards being significant in the CoLaus (β=-0.044, 95%CI -0.086 to -0.001, p=0.077).

**Figure 3.**
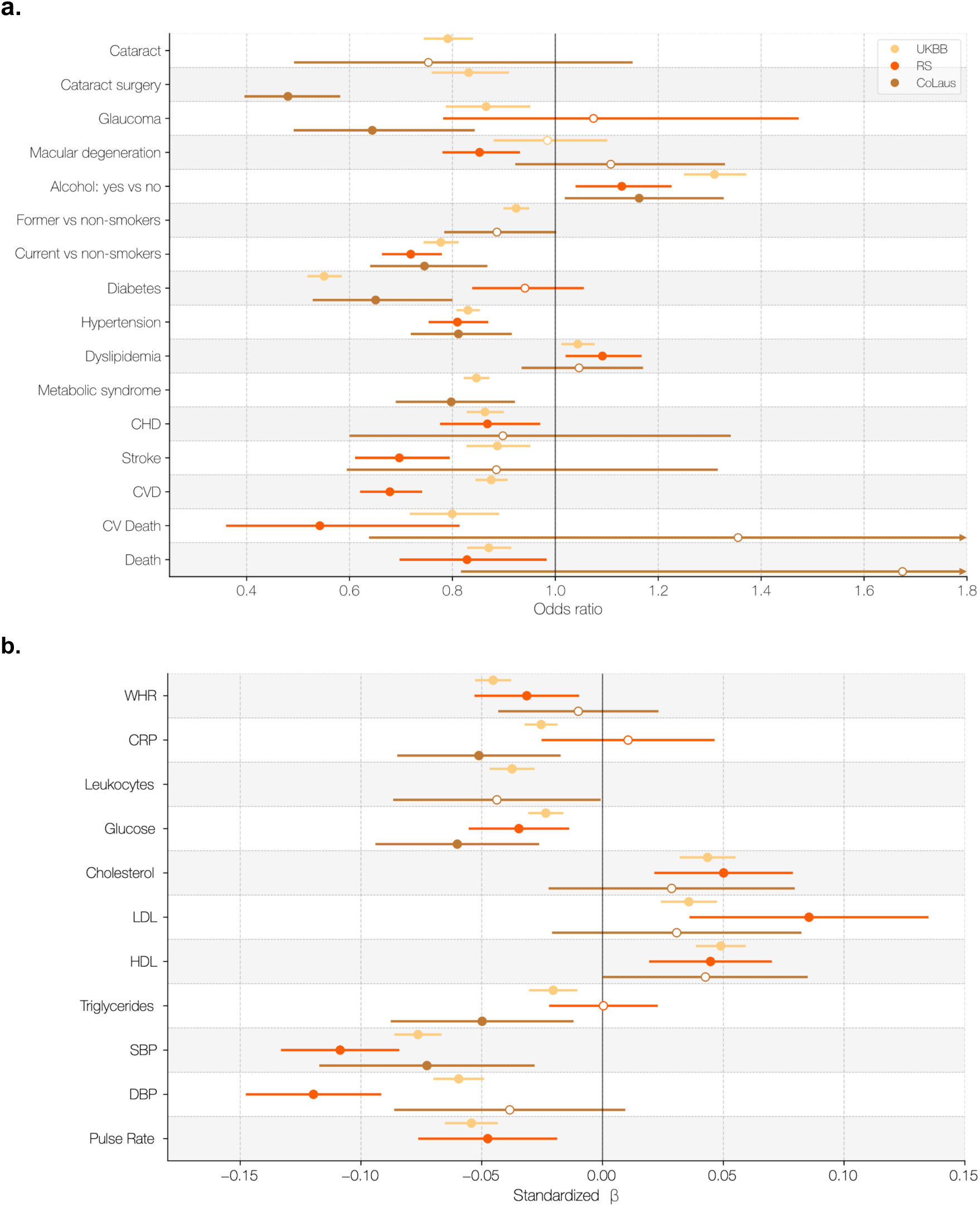
External validation of RetFit for the disease/risk factors association analyses in the Rotterdam Study and the CoLaus cohort. **a)** Standardized coefficients for RetFit from independent models predicting binary outcomes in three independent cohorts (UKBB, RS, and CoLaus). **b)** Standardized coefficients for RetFit and SETCRF from independent models predicting continuous outcomes. All models were corrected for sex, age, and BMI. Filled circles indicate significance after Benjamini-Hochberg correction (in both panels). UKBB: UK Biobank; RS: Rotterdam Study; CoLaus: CoLaus Study; CHD: Coronary Heart Disease; CVD: Cardiovascular Disease; CV Death: Cardiovascular Death; WHR: waist-to-hip ratio; CRP: C-reactive protein; LDL & HDL: low- and high-density lipoprotein; SBP & DBP: systolic and diastolic blood pressure.

### RetFit is an Independent Biomarker for Cardiovascular Diseases and Overall Mortality

To investigate RetFit’s prognostic power, we evaluated its ability to predict future cardiovascular events and all-cause mortality using Cox Proportional Hazard (PH) models and benchmarked the results to SETCRF in the UKBB. We fitted unadjusted and adjusted models (see Methods) to determine how RetFit and SETCRF predict disease independently and jointly with common risk factors. Moreover, we used LRT to determine whether RetFit or SETCRF was better at improving the predictive capacity of models. Finally, we replicated the analysis in the Rotterdam Study cohort.

In the UKBB, both RetFit and SETCRF were significantly prognostic for cardiovascular events before and after correcting for cardiovascular risk factors. However, when we added to the model known cardiovascular disease risk factors alongside sex and BMI, only RetFit remained significantly prognostic (RetFit: HR=0.936, 95%CI 0.903 to 0.970, p<0.001; SETCRF: HR=0.979, 95%CI 0.958 to 1.00, p=0.0653; see **Fig. 4a** and **Suppl. Fig. 2a** for the HR of CRF estimates expressed in METs). Interestingly, we found that adding SETCRF to a model with RetFit showed no significant improvement (LRT, 1 degree of freedom (df) *χ*^2^=2.68, p=0.102) while adding RetFit to the model with SETCRF did (LRT, 1 df *χ*^2^=12.2, p<0.001; see **Suppl. Table 5a**), suggesting that RetFit adds distinct cardiovascular disease information that SETCRF fails to capture. Kaplan-Meier curves demonstrated good stratification of increasing cardiovascular event risk with decreasing fitness score (both RetFit and SETCRF; **Fig. 4b** and **Suppl. Fig. 2b**) and, in the fully adjusted Cox model, the third, fourth, and fifth RetFit quintiles were significantly associated with fewer cardiovascular events, while for SETCRF, none were (**Fig. 4c** and **Suppl. Fig. 2c**). When both biomarkers were included in the fully adjusted model, only RetFit quintiles remained significantly associated with cardiovascular endpoints. Adding RetFit quintiles to a model with SETCRF quintiles improved the model (LRT, 4 df *χ*^2^=13.4, p<0.01) while the opposite was not true (LRT, 4 df *χ*^2^=4.78, p=0.311; see **Suppl. Table 6a**), suggesting, once again, that RetFit captured prognostic information not accounted for by SETCRF. RetFit’s prognostic power for cardiovascular events was corroborated in the Rotterdam Study (RS) cohort, where we found it was significantly associated with such an endpoint before (HR=0.668, 95%CI 0.617 to 0.723, p<0.001) and after adjusting for all the risk factors (HR=0.813, 95%CI 0.733 to 0.901, p<0.001). RetFit’s generalisation capabilities were further confirmed in the RS group analysis, where KM curves showed good stratification (**Fig. 4d**; **Suppl. Table 9** and **Suppl. Fig. 3a**) and, in the Cox analysis, all of the quintiles but the second were significantly associated with cardiovascular events before (HR as low as 0.306, 95%CI 0.221 to 0.424, p<0.001; see **Suppl. Fig. 3b–c**) and after adjusting for all the risk factors (HR as low as 0.608, 95%CI 0.420 to 0.880, p=0.00837; see **Fig. 4d**).

**Figure 4.**
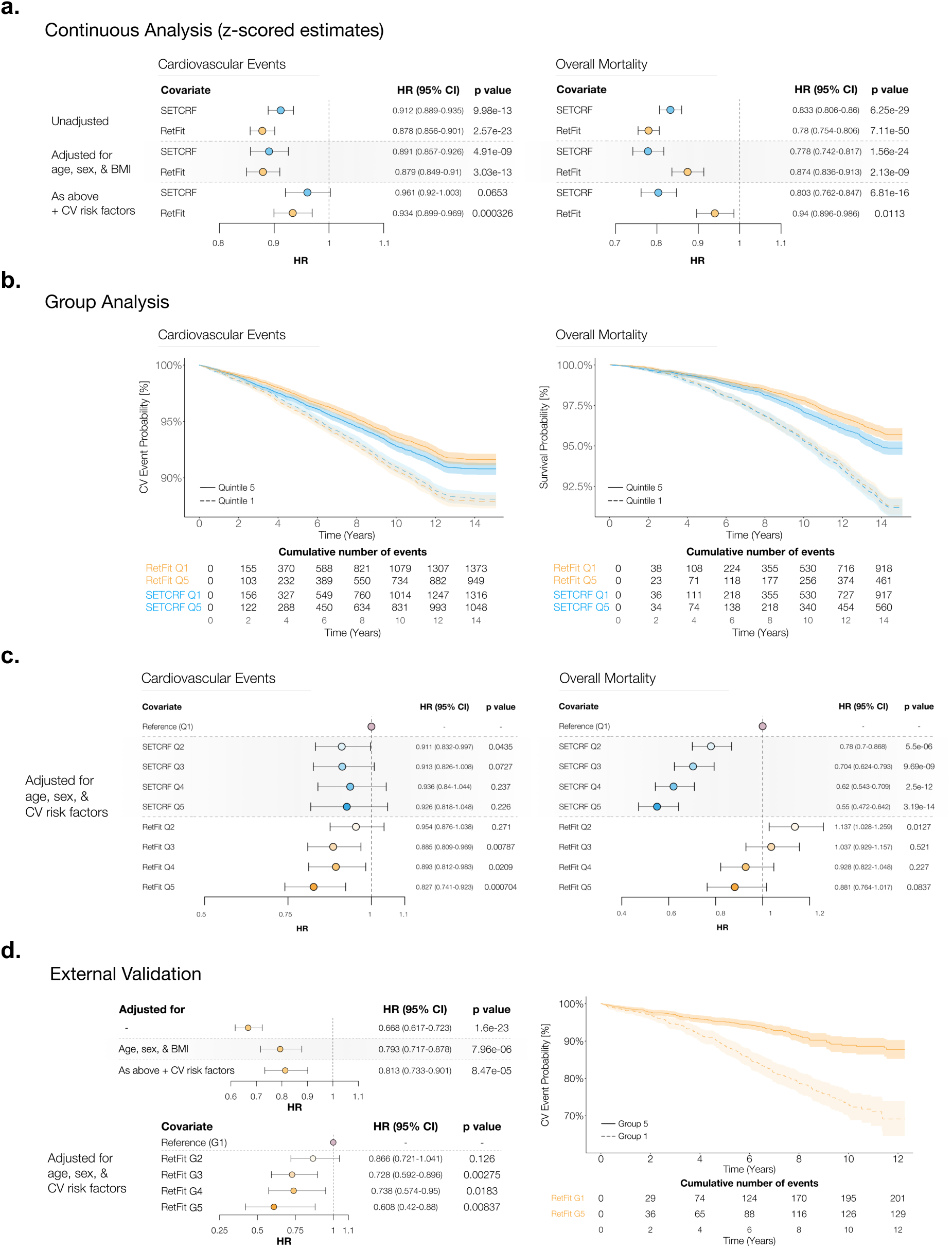
Prognostic performance and external validation of RetFit. **a)** Forest plots of the CRF estimates as a continuous parameter for cardiovascular events (left) and overall mortality (right) in the UKBB. **b)** Kaplan-Meier survival analysis of the RetFit and SETCRF (quintiles 1 and 5), and **c)** forest plots for all the quintiles in the UKBB. **d)** External validation of RetFit in the Rotterdam Study cohort. On the left side, from top to bottom: continuous analysis (z-scored) and group analysis. On the right side, Kaplan-Meier survival analysis (groups 1 and 5). For the group analysis, all the group thresholds were computed independently on the UKBB. UKBB: UK Biobank; HR: hazard ratio; CI: confidence interval; CV: cardiovascular.

When using overall mortality as the outcome in a continuous univariate setting, both RetFit and SETCRF were again significantly prognostic before and after correcting for cardiovascular risk factors (**Fig. 4a**; see **Suppl. Fig. 2a** for the HR of CRF estimates expressed in METs). When the estimates were included alongside all covariates, both significantly improved model fit (see **Suppl. Table 5b**). In the Kaplan-Meier analysis, we observed good stratification power (**Fig. 4b** and **Suppl. Fig. 2b**), while in the Cox analysis RetFit’s performance degraded after adjusting for demographics and the other cardiovascular risk factors (**Fig. 4c and Suppl. Fig. 2c**). Adding RetFit quintiles to a (fully adjusted) model with SETCRF quintiles significantly improved the model (LRT, 4 degrees of freedom *χ*^2^=19.7, p<0.001), and so did adding SETCRF quintiles to a model with RetFit quintiles (LRT, 4 degrees of freedom *χ*^2^=62.2, p<0.001) (see **Suppl. Table 6b**). Surprisingly, the second RetFit quintile showed an increased risk compared to the baseline, suggesting confounding effects might be at play. Additional details on the models’ fits are available in **Suppl. Tables 7** and **8**.

### RetFit is Genetically Distinct from SETCRF

To understand whether RetFit reveals new genes that may be important for CRF, we performed Genome-Wide Association Studies (GWAS) on both RetFit and SETCRF (see **Suppl. Fig. 4a–b** for genome-wide SNPs Manhattan plots). We found that RetFit was associated with 41 genes and SETCRF was associated with 25 genes. No significant genes were shared between RetFit and SETCRFs (**Fig. 5**; see **Suppl. Tables 11a** and **11b** for full list of genes). We also performed pathway analyses using the gene scores and found that RetFit was associated with 39 pathways and SETCRF was associated with 158 pathways. The significant pathways found for RetFit were mostly related to abnormalities of the eye, cell development mechanisms, and the nervous system. The 158 significant pathways we found for SETCRF were mostly associated with the cardiovascular system (see **Tables 1a** and **1b** for the 10 most significant pathways, **Suppl. Tables 12a–b** for their description, and **Suppl. Tables 12c–d** for the full list of significant pathways). Heritability (h^2^) estimates were similar for both traits (SETCRF h^2^=21.1%, SE=1%; RetFit h^2^=22.82%, SE=1%), and the genetic correlation of the two estimates was low (0.09, SE=0.03, p=0.008).

**Figure 5.**
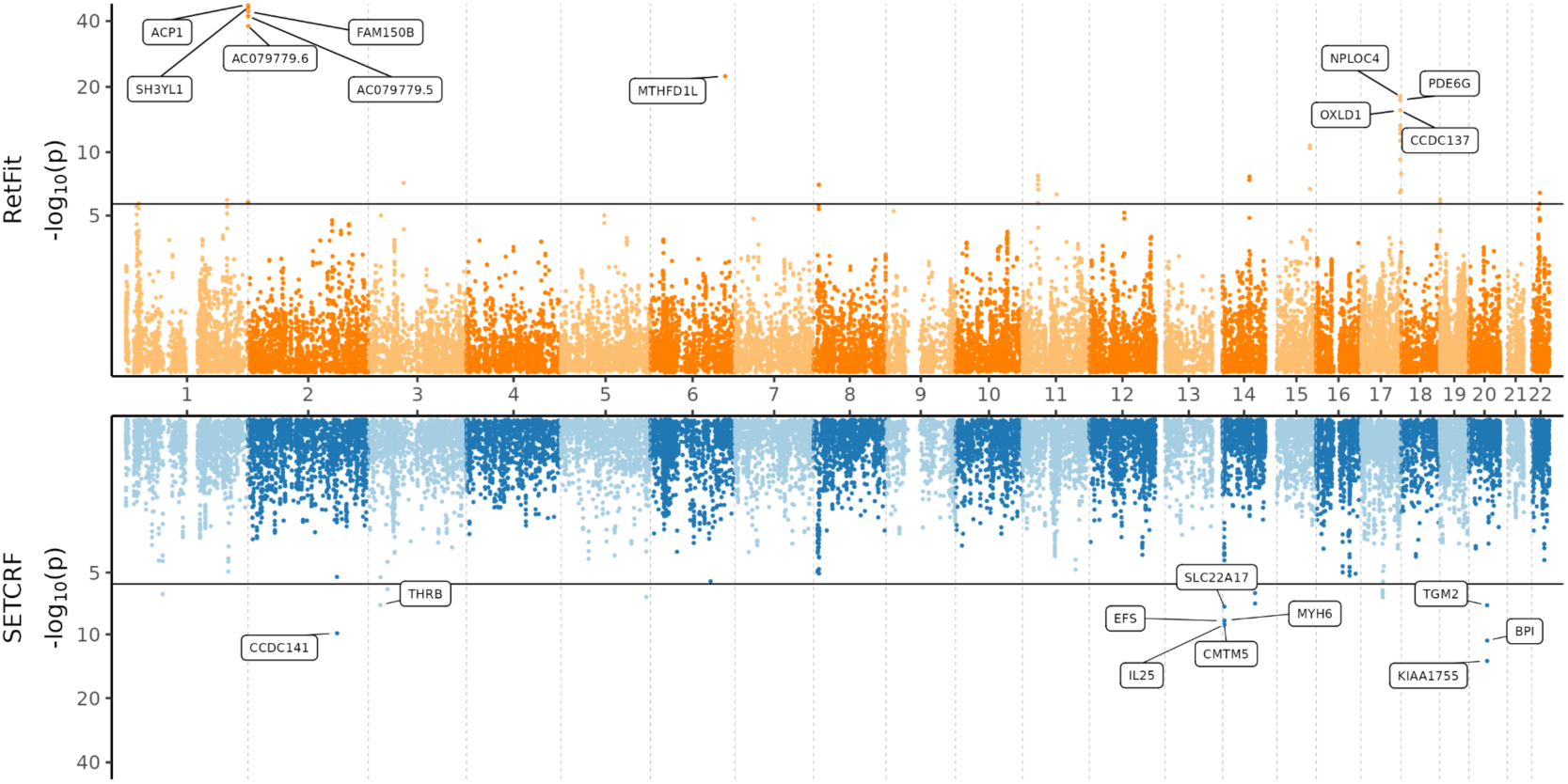
Gene enrichment analysis results for RetFit and SETCRF in the UKBB dataset. Miami plot showing all genes that were scored for the analysis using PascalX. Each dot corresponds to the p-value of one of the 24 922 genes scored. The x-axis shows the position of the gene in the genome, and the y-axis shows the significance of every gene score after a -log_10_ transformation, plotted on a log_2_ scale. The black line represents the significance threshold after Bonferroni correction. The upper panel shows the gene scores for RetFit, and the lower panel shows the gene scores for SETCRF. The 10 most significant genes are labeled for each trait.

**Table 1.**
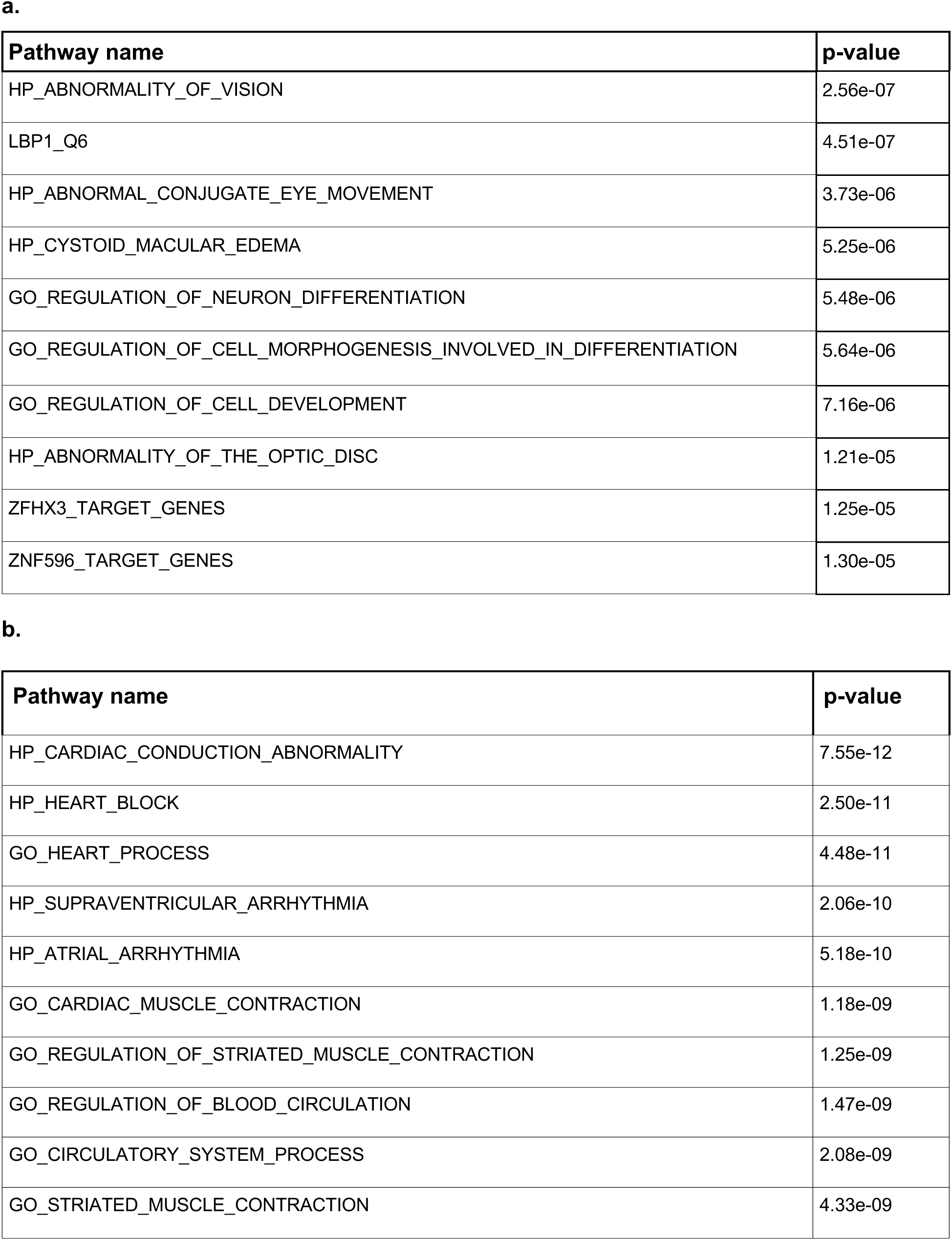
Pathway analysis of the two CRF estimates (UKBB dataset). List of 10 most significant pathways for **a)** RetFit and **b)** SETCRF. All the pathways available in MSIgDB v7.2 were scored using PascalX.

### Retinal Vasculature Associations with CRF

To better understand how RetFit might be predicting CRF from CFIs, we generated attention maps for RetFit to identify which image features are driving the CRF estimation. These attention maps qualitatively indicated that the model focused on the areas of the CFIs where the superior vascular beds typically lie (**Fig. 6a**). Furthermore, we used a state-of-the-art pipeline to extract quantitative vascular features from CFIs (hereafter referred to as Tangible Image Features, or TIFs) to assess their associations with both RetFit and SETCRF^28^. When uncorrected for covariates, all TIFs but three were associated with either RetFit or SETCRF (the Ratio tortuosity, the A diameter std, and the A median diameter for SETCRF and the A diameter std, the ratio median diameter and the ratio central retinal equivalent for RetFit; **Suppl. Fig. 5**). The number of arterial (A) bifurcations demonstrated the strongest correlation (Pearson *r* Coefficient) with both RetFit (r=0.259, 95%CI 0.251 to 0.267, p<0.001) and SETCRF (r=0.155, 95%CI 0.147 to 0.163, p<0.001). After accounting for age, sex, and BMI, 11 of the 18 TIFs remained associated with SETCRF, whereas 13 of the 18 TIFs were still associated with RetFit. (**Fig. 6b**). The number of A bifurcations remained the vascular property showing the strongest association with RetFit (β=0.287, 95%CI 0.263 to 0.311, p<0.001), but the vascular density ratio (arteries/veins) showed the strongest association with SETCRF (β=0.391, 95%CI 0.352 to 0.43, p<0.001) (**Fig. 6b**).

**Figure 6.**
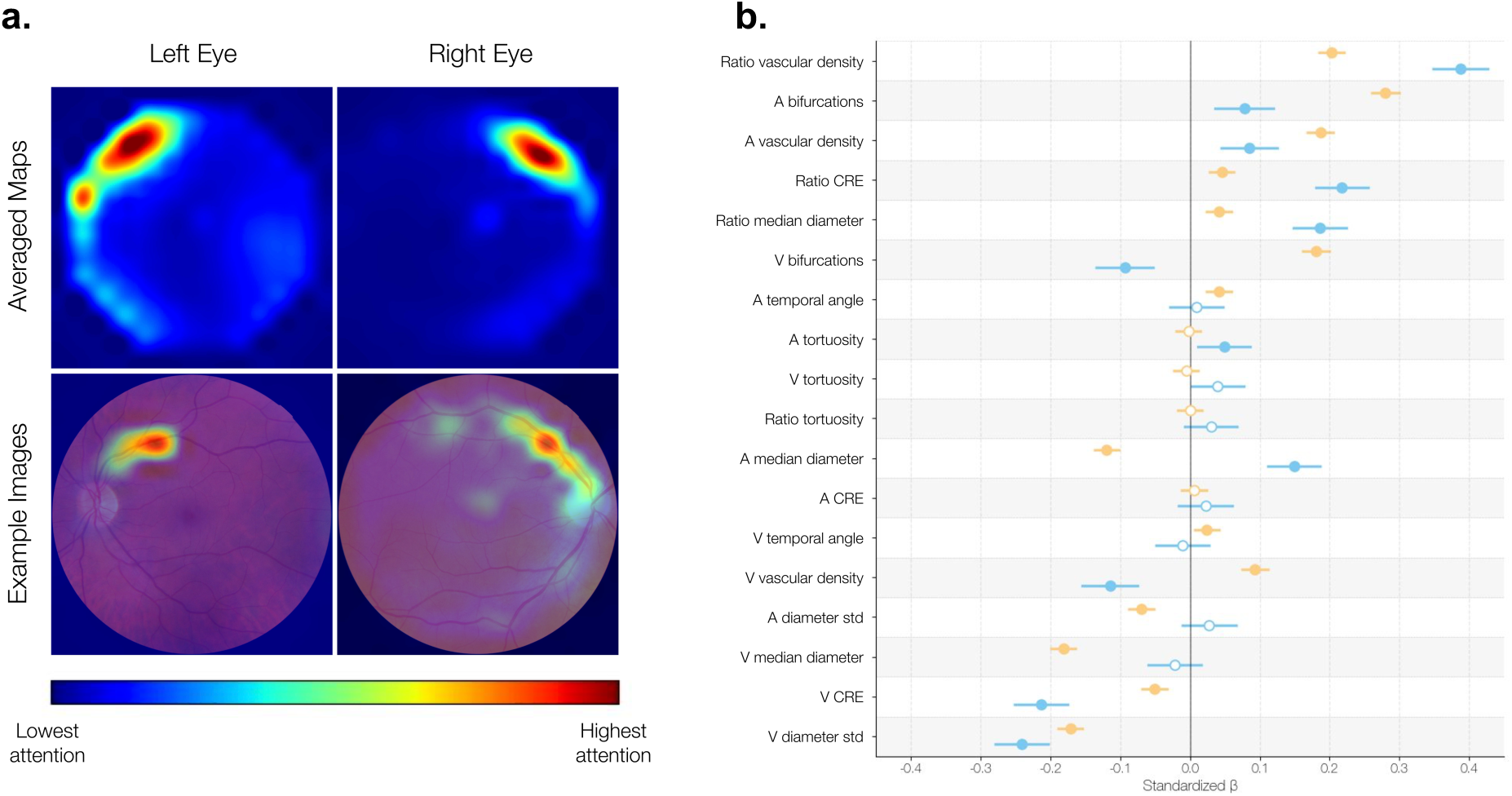
Attention mapping of the RetFit vision transformer model and association of RetFit and SETCRF with retinal vasculature features in the UKBB. **a.** The top two images show the average attention maps across all images from the UKBB for the left and right eyes, respectively. The bottom two images present examples of attention maps overlaid on CFIs for the left and right eye, providing a clear illustration of the specific regions the model focuses on when making predictions. **b.** Forest plot showing associations between retinal vascular features extracted from CFIs using VascX models^28^ and RetFit and SETCRF corrected for age, sex, and BMI. A: arteries; V: veins; CRE: central retinal equivalent; std: standard deviation.

## Discussion

Cardiorespiratory fitness (CRF) is among the most robust predictors of disease outcomes^1–6^, yet it remains challenging to measure accurately at scale^9–12^. In this study, we aimed to assess whether SETCRF (a submaximal exercise test to estimate CRF) could be reliably estimated from CFIs and to determine whether this estimator (that we call *RetFit)* correlates with current disease diagnoses and can predict future cardiovascular events. We found that, while RetFit only captures a moderate fraction of SETCRF’s variability (R^2^=0.2), it nevertheless provided diagnostic and prognostic information regarding disease, equaling the performance of direct SETCRF estimates reported in the literature^27^ and, in some cases, surpassing them. Interestingly, our GWAS conducted on both SETCRF and RetFit revealed no overlapping genetic associations, indicating distinct genetic underpinnings for these two fitness estimates. These findings highlight both the potential and limitations of retinal imaging as a surrogate measure for CRF, underscoring the need for continued exploration into the biological mechanisms linking retinal vascular characteristics with systemic health.

The prognostic power of objectively measured VO₂max in predicting future cardiovascular events is extensively documented across large-scale cohort studies^1–3,8,13^. A strong, inverse, and graded association exists between an individual’s VO₂max and the incidence of coronary artery disease, myocardial infarction, stroke, and cardiovascular mortality in general^3,29^. Previous work has demonstrated that CRF remains a significant predictor of cardiovascular events after adjustment for established multivariable risk scores, such as the Framingham Risk Score, or major cardiovascular risk factors (such as sex, age, and blood pressure)^1,30,31^. In our analysis, we made a similar observation for RetFit in the UKBB (i.e., it remained significant after adjustment for age, sex, BMI, SBP, HDL cholesterol, total cholesterol, smoking status, diabetes diagnosis, hypertension treatment, and previous cardiovascular events), but, interestingly, not for SETCRF. This finding may reflect fundamental differences between the two estimates: whereas SETCRF is derived from exercise performance, which is closely linked to established cardiovascular risk factors, RetFit leverages retinal image features that may quantify additional, subclinical markers of vascular health not captured by conventional measurements. Consequently, RetFit could complement existing risk scores, offering additional diagnostic and prognostic value to routinely acquired clinical data. It is worth noting that, in the study proposing SETCRF, Gonzales et al. (2021) reported that the estimate remained a significant predictor of cardiovascular events after adjustment for several risk factors^24^. In our analysis, significance was not reached (p=0.065) after adjusting for a slightly different set of covariates (including systolic blood pressure, HDL, and total cholesterol, and excluding several covariates from Gonzales et al.) and using a different definition of prior cardiovascular events. Differences in sample size (57 588 vs. 79 981, as we selected the subset of participants with retinal images, see **Suppl. Fig. 1**) and grouping strategy (quintiles vs. 1-MET increments) likely contributed to this discrepancy.

When we looked at disease associations, RetFit consistently showed a comparable diagnostic power to SETCRF across a broad range of analyses, with standardized beta coefficients and odds ratios often differing in magnitude but pointing in the same direction, underscoring its clinical relevance. We only noticed a major difference between the association of the two traits with a few traits, including cognitive performance metrics such as reasoning or prospective memory. In fact, the latter appear to be better captured by RetFit than SETCRF, pointing to potential systemic relevance beyond cardiovascular outcomes. Interestingly, the standardized effect size for LDL and total cholesterol differed in sign between RetFit and SETCRF, potentially reflecting the *cholesterol paradox,* where some studies have reported higher survival among older individuals with high levels of LDL and total cholesterol, possibly due to reverse causation or U-shaped associations^32–34^. As we previously discussed, RetFit showed superior prognostic power, independent of cardiovascular risk factors (e.g., HDL, total cholesterol, and SBP) for cardiovascular events. This was consistent with the results of the association analysis, where RetFit, after adjusting for age, sex, BMI, and socioeconomic status, showed a weaker correlation with SBP, DBP, HDL, LDL, total cholesterol, and triglycerides compared to SETCRF, indicating that RetFit is less tied to these factors. Beyond cardiovascular outcomes, RetFit also predicted overall mortality independently of conventional risk factors, underlining its broad clinical significance. We confirmed RetFit’s diagnostic and prognostic capabilities in two external and independent cohorts, namely, the Rotterdam Study and the CoLaus study. Besides validating the generalizability of our CFI-based fitness estimate beyond the UK Biobank population, this is particularly significant because it confirms RetFit’s potential as a highly scalable biomarker for CRF estimation in populations where maximal oxygen consumption or even submaximal exercise testing data are unavailable. Indeed, many large-scale population and clinical cohorts lack resource-intensive CRF measurements, but include CFI, in which case RetFit provides a novel, externally validated, and non-invasive method to retrospectively and prospectively estimate a robust, disease-relevant CRF-related measure, unlocking new opportunities for large-scale cardiovascular risk stratification and investigation of the molecular underpinnings of systemic health.

From the genetic analysis, we found that both RetFit and SETCRF yielded significant genetic associations; however, the genes and pathways identified were distinct, with no overlap observed between the two traits. The most significant hit we identified for RetFit, *ACP1* (p=5.84e-48), is a protein-encoding gene regulating cellular signaling by removing phosphate groups from tyrosine residues on proteins, known to be involved in coronary artery disease^35^ (see **Supp. Tables 10a** and **10b**). For SETCRF, our findings indicate *MYH6* (p=2.63e–09) as one of the strongest hits; this gene encodes the alpha-myosin heavy chain, the most abundant protein in the heart and a central determinant of cardiac muscle contraction, with established associations to cardiomyopathy^36^. Furthermore, we observed significant but distinct gene hits related to mitochondrial processes for both traits. In the case of RetFit, our analysis revealed a significant association with *MTHFD1L* (p=5.6e-23), a protein-encoding gene involved in the synthesis of tetrahydrofolate (THF) in the mitochondrion^37^. For SETCRF, we found a significant association with several genes such as *IL25* (p=1.05e-09), a cytokine promoting mitochondrial respiration^38^, *TGM2* (p=6.04e-08), an enzyme regulating mitophagy^39^, or *THRB* (p=5.91e-08), a thyroid hormone receptor, known to be involved with the mitochondria^40^. Furthermore, we identified several significant pathways linking SETCRF with CVD when running the pathway analysis (e.g., abnormality of cardiac conduction or impaired conduction of cardiac impulse, see **Supp. Table 11b** for pathway description), but not for RetFit, where pathways related to the cardiovascular system failed to reach significance. However, as we showed in the survival analysis, RetFit demonstrated better prognostic power for CV events than SETCRF, highlighting the need to better understand the differentiation between diagnostic and prognostic leverage and genetic mechanisms. In contrast, we found that the disease and genetic analyses seem to converge on endpoints associated with neuronal development, specifically, reasoning, visual memory, reaction time, and prospective memory. RetFit showed significant associations with each of these risk factors (whereas SETCRF did not), and multiple enriched pathways suggest that its genetic architecture influences processes related to the nervous system, neuronal development, and cerebral function (e.g., regulation of neuron differentiation, see **Supp. Table 11a**). Consequently, the associations we observed with cognitive markers are consistent with the genetic evidence, supporting the hypothesis that RetFit is functionally related to neuronal processes.

The physiological limit of CRF is fundamentally determined by the cardiovascular system’s ability to transport oxygen to metabolically active tissues^47^. While cardiac output and arterial oxygen content are critical components, CRF may also hinge on the integrity and function of the systemic vasculature, where oxygen exchange occurs^48^. The retinal circulation provides a unique, non-invasive window to observe this vasculature network in vivo, and morphological changes within it are well-established markers for systemic vascular pathology^49^. For example, narrowing of retinal arterioles and widening of venules, often summarized as a reduced arteriolar-to-venular ratio (AVR), are robustly associated with hypertension and the long-term risk of coronary heart disease^50^. Beyond vessel geometry, the branching complexity of the vascular tree, quantified by its fractal dimension, reflects the efficiency of tissue perfusion; a sparser, less complex network (lower fractal dimension) is linked to vascular rarefaction and has been shown to predict adverse outcomes like stroke^51^. Because these retinal features capture systemic vascular health, they may also reflect physiological pathways that underlie CRF. In line with this observation, most TIFs showed significant associations with both RetFit and SETCRF, with some of them displaying stronger associations with SETCRF than RetFit, despite the latter being based on CFIs.

It is critical to interpret these findings in the context of the study cohorts. We developed RetFit on a subset of the UKBB, which consists largely of individuals of White British ancestry, aged from 40 to 70. As a volunteer-based cohort, the UKBB is subject to healthy-volunteer and socio-economic selection biases: participants are typically healthier, less socioeconomically deprived, and report fewer health conditions than the general population^52^. However, the UKBB remains the largest and one of the only cohorts for which both CFIs and CRF estimates are available. In this regard, it is important to acknowledge that RetFit was modeled after SETCRF, which is itself a CRF estimate^24^. Notably, VO₂max was not objectively measured in any of the UKBB participants, and the validation cohort demonstrated higher exercise capacity than the average UKBB participant^24^. Such factors likely affect the accuracy of CRF estimation and, consequently, may also influence our analyses. It is also important to note that, since RetFit is derived from a deep learning (DL) pipeline, its distribution may exhibit a reduced standard deviation compared to other CRF estimates (due to the tendency of DL models to produce outputs that regress toward the mean, thereby compressing the range of predicted values^53^), and therefore yield inflated HRs when the latter are reported in units of MET. Given this, we opted to report the HR in units of standard deviation rather than METs whenever comparing the predictive power of RetFit and SETCRF in a continuous setting. We nevertheless report the HR in units of MET in the supplementary material to allow for a direct comparison with other DL and non-DL derived markers from the literature. Finally, the external validation cohorts (i.e., the Rotterdam Study and the CoLaus study) are also largely composed of individuals of White European ancestry.

In summary, in this work, we demonstrated that DL can generate a powerful and scalable biomarker from retinal images that reflects an individual’s health and disease risk. While conceived as a surrogate for CRF, our findings reveal a more complex and compelling story: RetFit’s superior prognostic power for future cardiovascular diseases, its comparable diagnostic capacity regarding disease outcomes and risk factors, combined with its distinct genetic architecture from SETCRF, establishes it not as a mere proxy but as a novel biological indicator. This reinforces the idea that the retinal microvasculature provides a unique window into a dimension of systemic health that is related to, but not fully captured by, traditional physiological fitness. As a purely image-derived estimate of CRF, RetFit offers a major practical advantage as it is much less invasive than classical cardiorespiratory fitness tests (i.e., VO_2_max test), which require expensive equipment, trained personnel, and maximal physical effort. Thus, RetFit can offer a more scalable alternative, in particular for frail and elderly populations, for whom CFIs often exist already or can be readily acquired, making it a promising tool for widespread clinical and population health use.

## Methods

### Data and quality control

The UK Biobank (UKBB) is a population-based cohort comprising approximately 488 000 participants, featuring extensive longitudinal phenotypic data, including comprehensive medical histories, with a median follow-up period of 10 years^54^. Cardiorespiratory fitness (CRF) was defined as maximal oxygen consumption or VO_2_max and estimated from heart rate responses obtained during submaximal exercise testing^24^. Standard 45° retinal color fundus images (CFIs) were acquired using a Topcon 3D-OCT 1000 Mark II imaging system.

For the present analysis, CRF estimates for 82 850 individuals measured as stated in^24^ were used. Additionally, a total of 173 814 CFIs from 84 813 individuals were accessible for analysis. Given our objective of evaluating the predictive capability of CFIs for estimating CRF, we implemented a previously published quality control (QC) procedure to ensure image integrity^55^. Specifically, image quality was initially quantified by professional graders on a subset of 1 000 CFIs, after which a convolutional neural network (CNN) was trained to replicate this human quality assessment. The top 75% highest-quality images, as determined by the CNN, were included in subsequent analyses. Since CRF and CFIs could be acquired at either instance 0 or instance 1, we matched each CFI with the corresponding CRF measurement from the same instance, prioritizing instance 0 and using instance 1 only if instance 0 data were unavailable. In total, we had matching CFI and CRF for 57 588 individuals (see **Suppl. Fig. 1**).

### Predicting CRF with RETFound

We utilized RETFound^22^, a novel foundation model for CFIs that leverages Google’s vision transformer large patch 16 architecture^56^. For predicting CRF, we modified some of the base hyperparameters established in the original RETFound manuscript^22^ and trained the model over 50 epochs, with a “warm-up” in the 10 first epochs (with the learning rate *r* monotonically increasing from 0 to 5 × 10^−4^), followed by a cosine annealing schedule (where *r* decreases from 5 × 10^−4^ to 1 × 10^−6^). We set the batch size to 16, the dropout rate to 0.4, and the image size to 224×224 pixels. After each epoch, we evaluated the model on a validation dataset and then kept the model that had the lowest mean square error across all epochs. To ensure robust evaluation, we implemented a 5-fold cross-validation strategy with a leave-one-out design: for each fold, the model was trained on 80% of the dataset and evaluated on the remaining left-out split. All the results we report in the manuscript were obtained by aggregating the predictions across the five left-out partitions. In addition, we fine-tuned separate models for each laterality, yielding five cross-validated models for the right eye and five for the left eye. To obtain the final subject- and instance-level prediction, we averaged the outputs from the left and right eyes when both images passed quality control, or used the single available prediction otherwise. Pearson correlations, R^2^, MAE, and RMSE were computed on the CRF averaged over both eyes.

### Disease Association

For the disease association analyses, we included a set of disease and general systemic risk factors that had either a binary or a continuous outcome known to affect disease susceptibility. We collected individual-level disease and risk factor information from the UKBB. We prioritized information provided through health-related records (Category 100091), biological samples (Category 100078), and Physical measures (Category 100006) over those obtained through the assessment center Touchscreen questionnaire (Category 100025) or verbal interviews (Category 100071), as self-reported measures are often unreliable due to high reporting errors or inconsistencies^57,58^. We made exceptions for alcohol intake and smoking status (i.e., never, previous, current). As studies have shown the prevalence of undiagnosed diabetic or pre-diabetic conditions and associated health complications^59,60^, we determined diabetes status using a combination of variables: an official diagnosis based on the date E10 first reported (insulin-dependent diabetes mellitus), HbA1C readings of larger or equal to 48 mmol/mol, or glucose levels of larger or equal to 11.1 mmol/l^61,62^. For participants who took antihypertensive medication, we added 10 mmHg to SBP and 5 mmHg to DBP values, and for those who took cholesterol-lowering medication, we added 1.4 mmol/L to LDL, 0.4 mmol/L to triglycerides, 1.6 mmol/L to total cholesterol, and subtracted 0.1 mmol/L from HDL. Participants who had total cholesterol levels higher than 5, HDL lower than 1, LDL higher than 4.1, triglycerides higher than 2.2, or took cholesterol-lowering medication were considered to have dyslipidemia. For blood pressure and pulse rate, we computed the average between (up to) the two automatic measurements per subject (taken one after the other, on the same visit). We defined hypertension as having SBP > 140 or DBP > 90 or the use of blood pressure medication. Following the International Diabetes Federation consensus^63^, we defined metabolic syndrome as having three or more of these phenotypes: large waist circumference (>= 80 cm for female or >= 94 cm for males), triglycerides > 1.7 or cholesterol medication, low HDL (< 1.29 for females or < 1.03 for males), high BP (SBP >= 130 and/or DBP >=85 and/or BP medication), and high glucose which we substituted with the above definition of diabetes since fasting glucose was not available. We defined cardiovascular (CV) death by reviewing all ICD-10-coded^64^ causes of death and including those corresponding to cardiovascular conditions^65,66^. All the fields used for the analysis and their descriptions can be found in **Suppl. Tables 3a–c**.

We used multiple linear regressions for continuous risk factors and multivariate logistic regressions for binary ones. All models were corrected for sex, age, BMI, income, educational level, and the Townsend deprivation index (the latter three forming the socioeconomic status). Furthermore, both the response and the independent variables that were used for the linear regressions were standardized. We also binarized all diseases collected as “date first diagnosed” for the logistic regressions. To determine the significance of regression coefficients, we computed p-values and corrected for multiple hypothesis testing using the Benjamini-Hochberg method^67^, using a false discovery rate of 0.05. We also used the log-likelihood ratio test (LRT) to compare the addition of independent information for RetFit and SETCRF by fitting a nested model (with either RetFit or SETCRF and the rest of the covariates) and a model with both predictors with the same set of covariates.

### Survival Analysis

We defined two distinct clinical endpoints for the survival analysis: cardiovascular events (obtained by aggregating Ischaemic Heart Disease (IHD), Myocardial Infarction (MI), stroke events, Non-Ischaemic Cardiomyopathies (NIC), and thrombotic events), and overall mortality. Time to event (TTE) was defined as the days between the date when the retinal image was taken and the date of the reported event. If the subject experienced multiple events, the first event after the imaging visit was used. We fitted Cox Proportional Hazard (PH) models with three levels of covariate adjustment to assess the robustness of the CRF estimates. First, we conducted univariate analyses including only the estimate of interest. Second, we adjusted for age, sex, and BMI, as these are key determinants of CRF. Finally, we further adjusted for additional cardiovascular risk factors that broadly matched those in the Framingham score^68^, i.e., age, sex, SBP, Total and HDL cholesterol, BMI, smoking status, diabetes diagnosis, and hypertension treatment (i.e., medication), plus whether the subject had a previous cardiovascular event. Hypertension treatment was defined by aggregating male- and female-specific medication and filtering for “blood pressure medication”. Diagnosis of previous cardiovascular events was included due to an increased risk of recurrence^69^. All the fields used for the analysis and their descriptions can be found in **Suppl. Tables 1–3**, while the distribution of events and continuous risk factors can be found in **Suppl. Table 4a.** For both estimates and for each clinical endpoint, we fit two independent models (i.e., one for RetFit and one for SETCRF) using continuous measures as predictors. Additionally, we conducted a group analysis by stratifying participants into quintiles based on their CRF estimates, reflecting a common practice in the literature where CRF estimates are often categorized into quantiles or fitness groups to assess dose-response relationships and facilitate clinical interpretation^11,6,70^. We applied the same stratification method to the RetFit score to compare its ability to stratify risk with that of the estimated CRF. To compare the different CRF estimates and investigate whether they improved the fit and the prognostic power of the different Cox models, we used the LRT to compare nested models (i.e., to compare a model with a fixed set of covariates and one of the estimates with a model with the same covariates and both estimates) (see **Suppl. Table 5–8**). To maximise the sample size while avoiding sparse data at longer follow-up times, we censored subjects at 15 years for both cardiovascular events and deaths.

### Genome-wide association study (GWAS)

GWAS were conducted using REGENIE^71^. Before performing the analyses, the genotypic data were processed following UKBB guidelines. Variants failing quality control thresholds (MAF < 0.01, MAC < 100, genotype missingness > 0.1, or Hardy-Weinberg equilibrium p < 1×10⁻¹⁵) were excluded. The GWAS were corrected for a set of covariates known to influence phenotypic variability (cardiovascular or eye-specific factors). The covariates were age at assessment center visit, sex, BMI, age-squared, sex-by-age, sex-by-age-squared, spherical power, cylindrical power, spherical power-squared, cylindrical power-squared, imaging instance, assessment center, genotype measurement batch, and the first 20 genetic Principal Components (PCs) (see **Suppl. Table 10** for corresponding UKBB data fields).

### Gene and Pathway Scoring

We computed gene and pathway scores using PascalX, a Python library specialized in gene and pathway scoring for GWAS summary statistics^72–74^. PascalX utilizes the sum of χ^2^ method to test for gene enrichment for gene scoring and a rank-based method for pathway scoring, which unifies the gene p-values distribution and aggregates it to χ^2^ distributed random variables. We used the UK10K panel for gene scoring, and scored both protein-coding genes and lincRNAs (long intergenic non-coding RNA) using the “saddle” method, taking into account all SNPs within a 50kb window around each gene and with a MAF of 0.05. We scored all pathways available in MSigDB v7.2 (n=31 120), fusing and rescoring any co-occurring genes located less than 100 kb apart. We adjusted all significant gene and pathway scores for multiple testing using the Bonferroni and Benjamini-Hochberg methods, respectively.

### Heritability and Genetic Correlation

We estimated SNP-based heritabilities for RetFit and SETCRF using the GCTA (Genome-wide complex trait analysis) software package^75^. We estimated the proportion of variance of the trait explained by the genetic data with the genomic-relatedness-based restricted maximum-likelihood (GREML) method^75,76^. We constructed genetic relationship matrices (GRM) for each chromosome using only directly genotyped SNPs with a MAF > 0.01 to reduce bias due to imputation quality and reduce computational burden. GRMs were then combined and used to estimate SNP-based heritability (h^2^) and genetic correlations (GC).

### Deriving Attention from RETFound

We generated attention maps from each of the five fine-tuned RETFound models to investigate which regions the model focuses on during prediction, and to gain insights into the representations learned by the model. Attention weights were extracted from the final attention block of the Vision Transformer, specifically capturing the softmax-normalized attention scores from the class token to the patch tokens (“CLS→patch attention”)^77^, resulting in 14 × 14 maps for each of the 16 attention heads. To account for the varying influence of individual attention heads, we performed a head sensitivity analysis. For each test image and each model fold, we ran 16 modified forward passes, each time muting a different attention head in the final block by zeroing the corresponding attention weights^78^. We used the squared absolute difference between the perturbed and unperturbed predictions as a proxy to quantify the head’s contribution. We then used these differences to compute a weighted average of the attention maps across heads, emphasizing the heads that most strongly influenced the prediction. We averaged the resulting head-weighted attention maps across all individuals and folds, separately for left and right eyes, to produce population-level maps. For visualization purposes only, we upsampled such maps from 14 × 14 to the original image resolution using bicubic interpolation.

### Vascular Associations

To explore the potential relationship between the retinal vasculature and both RetFit and CRF, we leveraged recently developed state-of-the-art deep learning models called *VascX* to extract 18 vascular traits from CFI^28^. *VascX* models work in a 2-step procedure, the first step being the segmentation of the CFI to separate retinal features (e.g., optic disc, vessels, and, more precisely, artery/vein), and the second step being the feature extraction from the masks that were output during the first step. In this case, we extracted 18 Tangible Image Features (TIFs) derived from the vessels’ segmentation masks, and they comprise features such as tortuosity or vessel density (see **Suppl. Table 13** for the full list) and were all separated between arteries and veins. For a more detailed description of the algorithms to compute these features, see Ortin-Vela et al.^19^ and Vargas Quiros et al.^19,28^. Once the TIFs were extracted, we used the same method to match them to RetFit/CRF estimates, meaning that values were averaged between eyes if both eyes were used to compute the average for RetFit. We then computed Pearson correlation coefficients and associated tests for each vascular trait independently. Multiple linear regressions with either Retfit or SETCRF as the response variable and each of the 18 TIFs as the dependent variable were also fitted, and all models were corrected for age, sex, and BMI. Multiple hypothesis testing was addressed using Benjamini-Hochberg, and all reported p-values have been corrected accordingly.

### External Validation

To investigate the generalizability of RetFit, we replicated the disease association and survival analyses in two different cohorts, namely the Rotterdam Study (RS) and the CoLaus cohort. We performed quality control on the CFIs in these two studies using the same QC method mentioned under the *Data and quality control* section. We used the model weights fine-tuned on UKBB data to predict RetFit for all the images from the two external cohorts deemed of good enough quality. Given the 5-fold cross-validation strategy we adopted for the development and validation of the model in the UKBB, we obtained 5 independent models for each laterality. Therefore, we predicted five values of RetFitfor each image, which we averaged to obtain the final estimate for each CFI. For the RS, a quality measure was available for every image, so we selected the single image per visit and per subject with the highest quality. For the CoLaus, when subjects had CFIs that passed QC for both lateralities, we further took the mean between the two RetFit values. In the RS, when subjects had images from multiple visits, we favored the instance with the highest proportion of non-missing values for the risk factors and diseases we analysed. In the CoLaus, as CFIs were not acquired at the same time as the risk factors and disease statuses, we selected the instance for the risk factors and disease that was the closest to the date of imaging. After this phase, every subject had only one CFI and one matching RetFit value that was further used for downstream analyses.

For the continuous risk factor and binary disease variables in the RS and the CoLaus, we fitted similar linear models (linear regression and logistic regression, respectively) as we did for the UKBB. As neither of these two cohorts measured cardiorespiratory fitness estimates, we focused on the replication of effect sizes and directionality of the associations, and on associations with outcomes that were not available in the UKBB. We defined dyslipidemia and corrected BP, cholesterol (total cholesterol, LDL, and HDL), and triglycerides as described under the *Disease Association* section. All models were corrected for sex, age, and BMI, and both independent variables and continuous response variables were standardized. We once again computed p-values and corrected for multiple hypothesis testing using the Benjamini-Hochberg method and using a false discovery rate of 0.05. The distribution of events and continuous risk factors for both the RS and the CoLaus can be found in **Suppl. Table 4b–c.**

For the external validation of RetFit in a survival analysis setting, we defined the “cardiovascular events” endpoint by aggregating coronary heart disease, heart failure events, and stroke events. The time to event was defined as the number of days between the date the retinal image was taken and the date of the reported event. Similarly to the analysis in the UKBB, we fitted Cox Proportional Hazard (PH) models with three levels of adjustment to assess the generalisability of RetFit independent of several risk factors. In the univariate analysis, we included only RetFit; in the second step of adjustment we included age, sex, and BMI; in the complete model, we further adjusted for additional cardiovascular risk factors that broadly matched those in the Framingham score^68^, i.e., age, sex, SBP, Total and HDL cholesterol, BMI, smoking status and hypertension treatment (i.e., medication). Different from the analysis in the UKBB dataset, we were not able to adjust for diabetes diagnosis due to such information being available for a very limited number of subjects. Furthermore, we did not include previous cardiovascular events as a risk factor, as the subjects in the Rotterdam study were cardiovascular-event-free at the time of enrollment. We defined the risk groups for the group analysis using the threshold we independently computed in the UKBB. Similarly to the UKBB, we limited the follow-up for cardiovascular events to 12 years to maximise sample size while avoiding sparse data at longer durations.

## Data Availability

Genome-wide association study (GWAS) summary statistics generated in this study will be made publicly available on Zenodo following peer-reviewed publication. Image-derived phenotypic data will be available exclusively through the UK Biobank cohort platform after publication, subject to restricted access. The analysis code supporting the findings of this study will be released on GitHub upon peer-reviewed publication of the manuscript (https://github.com/BergmannLab/).

https://www.ukbiobank.ac.uk/enable-your-research/apply-for-access

https://www.erasmusmc.nl/en/research/departments/epidemiology

https://www.colaus-psycolaus.ch/colaus

## Author Information

VascX Consortium: Michael Beyeler ^1,2^; Ciara Bergin ^3^; Sven Bergmann ^1,2,4^; Dennis Bontempi ^1,2^; Sacha Bors ^1,2^; Leah Böttger ^1,2^; Bogdan Draganski ^5,6,7^; Adham Elwakil ^3,8^; Györgyi V. Hamvas ^9,10^; Janna Hastings ^11,12^; Ilaria Iuliani ^1,2^; Caroline C.W. Klaver ^13,14,15,16^; Ihor Kuras ^6,7^; Bart Liefers ^13,14^; Ilenia Meloni ^3,8^; Sofia Ortin Vela ^1,2^; David Presby ^1,2^; Ian Quintas ^1,2^; José Vargas Quiros ^13,14,^; Marc Schindewolf ^9,10^; Reinier O. Schlingemann ^3,17^; Mattia Tomasoni ^3,8^; Olga Trofimova ^1,2^.

1. *Department of Computational Biology, University of Lausanne, Lausanne, Switzerland*
2. *Swiss Institute of Bioinformatics, Lausanne, Switzerland*
3. *Department of Ophthalmology, University of Lausanne, Fondation Asile des Aveugles, Jules Gonin Eye Hospital, Lausanne, Switzerland.*
4. *Department of Integrative Biomedical Sciences, University of Cape Town, Cape Town, South Africa*
5. *Department of Neurology, Max Planck Institute for Human Cognitive and Brain Sciences, Germany*
6. *Insel University Hospital, Bern, Switzerland*
7. *Department of Clinical Neuroscience, Lausanne University Hospital and University of Lausanne, Switzerland*
8. *Platform for Research in Ocular Imaging, Fondation Asile des Aveugles, Jules Gonin Eye Hospital, Lausanne, Switzerland.*
9. *Inselspital, Bern University Hospital, University of Bern, Switzerland.*
10. *Department for BioMedical Research, Bern University Hospital, University of Bern, Switzerland.*
11. *Institute for Implementation Science in Health Care, Faculty of Medicine, University of Zurich, Zürich, Switzerland*
12. *School of Medicine, University of St Gallen, St. Gallen, Switzerland*
13. *Department of Ophthalmology, Erasmus University Medical Center, Rotterdam, The Netherlands.*
14. *Department of Epidemiology, Erasmus University Medical Center, Rotterdam, The Netherlands.*
15. *Department of Ophthalmology, Radboud University Medical Center, Nijmegen, the Netherlands.*
16. *Institute of Molecular and Clinical Ophthalmology, University of Basel, Switzerland.*
17. *University Medical Centres, Amsterdam, The Netherlands.*

## Data and Code Availability

The UK Biobank data used in this study are protected and not publicly available due to data privacy regulations. Access to UK Biobank data is granted to eligible researchers through an application process administered by the UK Biobank management team, subject to approval and payment of applicable access fees. Permission to access and analyse the UK Biobank data for the present study was granted under application number 90947.

Genome-wide association study (GWAS) summary statistics generated in this study will be made publicly available on Zenodo following peer-reviewed publication. Image-derived phenotypic data will be available exclusively through the UK Biobank cohort platform after publication, subject to restricted access. The analysis code supporting the findings of this study will be released on GitHub upon peer-reviewed publication of the manuscript.

Data from the Rotterdam Study and CoLaus studies can be made available to researchers upon reasonable request through a data transfer agreement.

## Funding

This research was supported by the Swiss National Science Foundation grant no. CRSII5 209510 for the “VascX” Sinergia project.

## Competing Interest

All the authors declare no competing interests, including both financial and non-financial interests.

## Notes

### Competing Interest Statement

The authors have declared no competing interest.

### Clinical Trial

NL6645 / NTR6831

### Funding Statement

This work was supported by the Swiss National Science Foundation grant no. CRSII5 209510 for the "VascX" Sinergia project. The authors declare no conflicts of interest.

### Author Declarations

The UK Biobank data used in this study are protected and not publicly available due to data privacy regulations. Access to UK Biobank data is granted to eligible researchers through an application process administered by the UK Biobank management team, subject to approval and payment of applicable access fees. Permission to access and analyse the UK Biobank data for the present study was granted under application number 90947. The external validation data from the Rotterdam Study and the CoLaus Study can be made available to researchers upon reasonable request through a data transfer agreement. The Rotterdam Study has been approved by the Medical Ethics Committee of the Erasmus MC (registration number MEC 02.1015) and by the Dutch Ministry of Health, Welfare and Sport (Population Screening Act WBO, license number 1071272-159521-PG). The OphthalmoLaus study received ethics approval from the "Commission Cantonale d'Éthique de la Recherche sur l'Être Humain" under project number PB_2019-00168.

### Summary of Updates

External validation of the biomarker.

