## Supplementary Figures and Tables for "RetFit: A Novel Deep Learning-Based Biomarker of Cardiorespiratory Fitness Derived From the Retina"

**Affiliations |** [1] Department of Computational Biology**,** University of Lausanne, Lausanne, Switzerland. [2] Swiss Institute of Bioinformatics, Lausanne, Switzerland. [3] Department of Ophthalmology, Erasmus University Medical Center, Rotterdam, The Netherlands. [4] Department of Epidemiology, Erasmus University Medical Center, Rotterdam, The Netherlands. [5] Department of Ophthalmology, Radboud University Medical Center, Nijmegen, The Netherlands. [6] Institute of Molecular and Clinical Ophthalmology, University of Basel, Switzerland. [7] Department of Ophthalmology, University of Lausanne, Fondation Asile des Aveugles, Jules Gonin Eye Hospital, Lausanne, Switzerland. [8] Platform for Research in Ocular Imaging, Fondation Asile des Aveugles, Jules Gonin Eye Hospital, Lausanne, Switzerland. [9] Department of Integrative Biomedical Sciences, University of Cape Town, Cape Town, South Africa.

$: see Author information for full list of co-authors.

***: Corresponding author |** David Presby, Ph.D., Department of Computational Biology; Genopode, office 2025.1 - CH1015 Lausanne, Switzerland.

**Suppl. Table 1 | Data fields used to define cardiovascular endpoints for the survival analysis in the UK Biobank.**

| Event Category | UKB Data Field | Field Name |
|---|---|---|
| **IHD** (Ischaemic Heart Disease) | 131296 | Date angina first reported |
| | 131304 | Date other acute IHD first reported |
| | 131306 | Date IHD first reported |
| **MI** (Myocardial Infarction) | 131298 | Date acute MI first reported |
| | 131300 | Date subsequent MI first reported |
| | 131302 | Date complications after MI first reported |
| | 42000 | Date of myocardial infarction |
| **Stroke** | 42006 | Date of stroke |
| | 42008 | Date of ischaemic stroke |
| | 42010 | Date of intracerebral haemorrhage |
| **NIC** (Non-ischaemic Cardiomyopathies) | 131338 | Date cardiomyopathy first reported |
| | 131340 | Date cardiomyopathy in other diseases first reported |
| | 131288 | Date hypertensive heart disease first reported |
| | 131292 | Date hypertensive heart and renal disease first reported |
| **Thrombotic Event** | 131308 | Date pulmonary embolism first reported |
| | 131388 | Date arterial embolism and thrombosis first reported |
| | 131400 | Date other venous embolism and thrombosis first reported |

**Suppl. Table 2 | Data fields used to define risk factors for the survival analysis in the UK Biobank.**

| **Risk Factor** | **UKB Data Field** | **Field Name** |
|---|---|---|
| Age | 21003 | Age at assessment |
| Sex | 22001 | Sex |
| Systolic Blood Pressure | 4080 | Systolic blood pressure (automated reading) |
| Smoking status | 20116 | Smoking status |
| BMI | 21001 | BMI |
| Total Cholesterol | 30690 | Cholesterol |
| HDL Cholesterol | 30760 | HDL cholesterol |
| Diabetes | 2976 | Age diabetes diagnosed |
| | 30740 | Glucose |
| | 30750 | Glycated haemoglobin (HbA1c) |
| | 2443 | Diabetes diagnosed by doctor |
| Medicated for Hypertension | 6153 | Medication for cholesterol, blood pressure, diabetes, or take exogenous hormones |
| | 6177 | Medication for cholesterol, blood pressure or diabetes |
| Previous Cardiovascular Event | 131296, 131304, 131306, 131298, 131300, 131302, 42000, 42006, 42008, 42010, 131338, 131340, 131288, 131292 | - |

**Suppl. Table 3 | Data fields used for the disease association analysis in the UK Biobank.**
**a)** Covariates **b)** Continuous risk factors **c.** Binary outcomes**.**

**a.**

| Covariate | UKB Data Field |
|---|---|
| Age | 21022 |
| Sex | 22001 |
| BMI | 21001 |
| Average total household income before tax | 738 |
| Qualifications | 6138 |
| Townsend deprivation index | 22189 |

**b.**

| Endpoint | UKB Data Field |
|---|---|
| Reasoning | 20016 |
| HbA1c | 30750 |
| CRP | 30710 |
| IGF-1 | 30770 |
| Pack years of smoking | 20161 |
| Reaction time | 20023 |
| Glucose | 30740 |
| Cholesterol | 30690 |
| HDL | 30760 |
| LDL | 30780 |
| WHR (waist circumference/hip circumference) | 48 / 49 |
| Triglycerides | 30870 |
| Leukocytes | 30000 |
| Systolic Blood Pressure (SBP) | 4080 |
| Diastolic Blood Pressure (DBP) | 4079 |
| Pulse Rate | 102 |

**c.**

| **Endpoint** | **UKB Data Field** |
|---|---|
| Alcohol (yes/no) | 1558 |
| Cancer | 2453 |
| Eye problems or disorders (cataract, cataract surgery, glaucoma, macular degeneration) | 6148 |
| Diabetes | 2443, 2976 |
| Prospective memory | 20018 |
| Smoking (current/previous vs never) | 20116 |
| Visual memory | 399 |
| Atherosclerosis | 131380 |
| CHD | 131296, 131304, 131306 |
| MI | 131298, 131300, 131302, 42000 |
| Stroke | 42006, 42008, 42010 |
| CV Death | 40001 |
| Death | 40000 |

**Suppl. Table 4 | Clinical characteristics of the study cohorts.** Descriptive statistics for **a)** UK Biobank cohort (N=57 588), **b)** The Rotterdam Study (N=8 885), and **c)** The CoLaus cohort (N=2 260). To maximise the sample size while avoiding sparse data at longer follow-up times, we limited incident cardiovascular events and deaths to the first 15 years in the UK Biobank and to the first 12 years in the Rotterdam Study. Total Cholesterol and HDL are reported in mmol/L. BMI: body mass index; SBP: systolic blood pressure. CV: cardiovascular.

**a.**

| **Covariate** | **Mean (IQR or percentage)** |
|---|---|
| **Age** | 56.1 (13) |
| **Sex** | |
| **Male** | 26 942 (46.8%) |
| **Female** | 30 646 (53.2%) |
| **BMI** | 27 (5.47) |
| **SBP** | 136.2 (24) |
| **Total Cholesterol** | 5.72 (1.49) |
| **HDL** | 1.50 (0.517) |
| **Diabetes Diagnosis** | 2 747 (5%) |
| **Smoking Status** | |
| **Current** | 32 871 (8.2%) |
| **Former** | 19 726 (34.3%) |
| **Never** | 4 734 (57.1%) |
| **Prevalent CV events** | 8 438 (14.7%) |
| **Incident CV events (15 years)** | 6 141 (10.7%) |
| **Treated for hypertension** | 10 596 (18.4%) |
| **All-cause mortality (15 years)** | 3 831 (6.7%) |

**b.**

| Covariate | Mean (IQR or percentage) |
|---|---|
| **Age** | 65 (19.07) |
| **Sex** | |
| **Male** | 3843 (43.2%) |
| **Female** | 5042 (56.8%) |
| **BMI** | 27.6 (5.46) |
| **SBP** | 141.4 (32) |
| **Total Cholesterol** | 5.39 (1.5) |
| **HDL** | 1.48 (0.56) |
| **Diabetes Diagnosis** | 1023 (11.5%) |
| **Smoking Status** | |
| **Current** | 1475 (80%) |
| **Non-smoker** | 7108 (16.6%) |
| **Prevalent CV events** | 1639 (18.5%) |
| **Incident CV events (12 years)** | 855 (10%) |
| **Treated for hypertension** | 3309 (37.2%) |
| **All-cause mortality (12 years)** | 372 (4.2%) |

c.

| Covariate | Mean (IQR or percentage) |
|---|---|
| **Age** | 64.1 (14.0) |
| **Sex** | |
| **Male** | 1025 (45.4%) |
| **Female** | 1235 (54.6%) |
| **BMI** | 26.2 (5.82) |
| **SBP** | 129.4 (25.5) |
| **Total Cholesterol** | 5.27 (1.3) |
| **HDL** | 1.58 (0.5) |
| **Diabetes Diagnosis** | 184 (8.1%) |
| **Smoking Status** | |
| **Current** | 357 (16.2%) |
| **Previous** | 831 (36.8%) |
| **Non-smoker** | 917 (40.6%) |
| **Prevalent CV events** | - |
| **Incident CV events (12 years)** | - |
| **Treated for hypertension** | 671 (29.7%) |
| **All-cause mortality (12 years)** | 23 (1.0%) |

**Suppl. Table 5 | Log-likelihood ratio tests for Cox PH models (continuous analysis in the UK Biobank).** Here, “demographics” refers to the Cox PH models adjusted for age, sex, and BMI - while “demographics + CV” refers to those adjusted additionally for all the other cardiovascular risk factors (SBP, HDL cholesterol, total cholesterol, smoking status, diabetes diagnosis, hypertension treatment, and previous cardiovascular event(s)). **a)** Cox PH models with cardiovascular endpoints as outcome, and **b)** with overall mortality as outcome. df: degrees of freedom. CV: cardiovascular.

**a.**

| Model | Chi-square test statistics | df | p-value |
|---|---|---|---|
| **Adding RetFit to a model with Demographics + SETCRF** | 47.361 | 1 | 5.905e-12 |
| **Adding SETCRF to a model with Demographics + RetFit** | 28.136 | 1 | 1.131e-07 |
| **Adding RetFit to a model with Demographics + CV + SETCRF** | 12.232 | 1 | 0.0004697 |
| **Adding SETCRF to a model with Demographics + CV + RetFit** | 2.6794 | 1 | 0.1017 |

**b.**

| Model | Chi-square test statistics | df | p-value |
|---|---|---|---|
| **Adding RetFit to a model with Demographics + SETCRF** | 28.327 | 1 | 1.025e-07 |
| **Adding SETCRF to a model with Demographics + RetFit** | 95.971 | 1 | 2.2e-16 |
| **Adding RetFit to a model with Demographics + CV + SETCRF** | 4.4045 | 1 | 0.03584 |
| **Adding SETCRF to a model with Demographics + CV + RetFit** | 62.914 | 1 | 2.159e-15 |

**Suppl. Table 6 | Log-likelihood ratio tests for Cox PH models (group analysis in the UK Biobank).** Here, “demographics” refers to the Cox PH models adjusted for age, sex, and BMI - while “demographics + CV” refers to those adjusted additionally for all the other cardiovascular risk factors (SBP, HDL cholesterol, total cholesterol, smoking status, diabetes diagnosis, hypertension treatment, and previous cardiovascular event(s)). **a)** Cox PH models with cardiovascular endpoints as outcome, and **b)** with overall mortality as outcome. df: degrees of freedom; CV: cardiovascular; Q: quintiles.

**a.**

| Model | Chi-square test statistics | df | p-value |
|---|---|---|---|
| Adding RetFit Q to a model with Demographics + SETCRF Q | 47.365 | 4 | 1.28e-09 |
| Adding SETCRF Q to a model with Demographics + RetFit Q | 20.874 | 4 | 0.0003355 |
| Adding RetFit Q to a model with Demographics + CV + SETCRF Q | 13.441 | 4 | 0.009312 |
| Adding SETCRF Q to a model with Demographics + CV + RetFit Q | 4.7771 | 4 | 0.3109 |

**b.**

| Model | Chi-square test statistics | df | p-value |
|---|---|---|---|
| Adding RetFit Q to a model with Demographics + SETCRF Q | 42.526 | 4 | 1.298e-08 |
| Adding SETCRF Q to a model with Demographics + RetFit Q | 92.602 | 4 | 2.2e-16 |
| Adding RetFit Q to a model with Demographics + CV + SETCRF Q | 19.732 | 4 | 0.0005642 |
| Adding SETCRF Q to a model with Demographics + CV + RetFit Q | 62.186 | 4 | 1.007e-12 |

**Suppl. Table 7 | Comparison of the different Cox PH models (continuous analysis in the UK Biobank). a)** Cox PH models with cardiovascular endpoints as outcome, and **b)** with overall mortality as an outcome. Here, “demographics” refers to the Cox PH models adjusted for age, sex, and BMI - while “demographics + CV” refers to those adjusted additionally for all the other cardiovascular risk factors (SBP, HDL cholesterol, total cholesterol, smoking status, diabetes diagnosis, hypertension treatment, and previous cardiovascular event(s)). CI: concordance index; CV: cardiovascular; LL: model log-likelihood; AIC: Akaike Information Criterion.

**a.**

| Model | CI | LL-null model | LL-full model | AIC |
|---|---|---|---|---|
| **Demographics** | 0.6967168 | -66 860.64 | -65 346.33 | 130 698.7 |
| **+ RetFit** | 0.6982309 | | -65 319.58 | 130 647.2 |
| **+ SETCRF** | 0.6978317 | | -65 329.19 | 130 666.4 |
| **+ RetFit + SETCRF** | 0.6991379 | | -65 305.51 | 130 621.0 |
| **Demographics + CV** | 0.7207364 | -57 211.09 | -55 503.21 | 111 028.4 |
| **+ RetFit** | 0.7209280 | | -55 496.74 | 111 017.5 |
| **+ SETCRF** | 0.7208950 | | -55 501.51 | 111 027.0 |
| **+ RetFit + SETCRF** | 0.7210542 | | -55 495.40 | 111 016.8 |

**b.**

| Model | CI | LL null model | LL full model | AIC |
|---|---|---|---|---|
| **Demographics** | 0.7182870 | -42 689.65 | -41 460.70 | 82 927.39 |
| **+ RetFit** | 0.7198590 | | -41 442.64 | 82 893.28 |
| **+ SETCRF** | 0.7217663 | | -41 408.82 | 82 825.64 |
| **+ RetFit + SETCRF** | 0.7228985 | | -41 394.65 | 82 799.31 |
| **Demographics + CV** | 0.7367081 | -36 356.86 | -35 087.58 | 70 197.17 |
| **+ RetFit** | 0.7370390 | | -35 084.36 | 70 192.73 |
| **+ SETCRF** | 0.7385669 | | -35 055.11 | 70 134.22 |
| **+ RetFit + SETCRF** | 0.7387833 | | -35 052.91 | 70 131.81 |

**Suppl. Table 8 | Comparison of the different Cox PH models (group analysis in the UK Biobank). a)** Cox PH models with cardiovascular endpoints as outcome, and **b)** with overall mortality as an outcome. Here, "demographics" refers to the Cox PH models adjusted for age, sex, and BMI - while "demographics + CV" refers to those adjusted additionally for all the other cardiovascular risk factors (SBP, HDL cholesterol, total cholesterol, smoking status, diabetes diagnosis, hypertension treatment, and previous cardiovascular event(s)). CI: concordance index; CV: cardiovascular; LL: model log-likelihood; AIC: Akaike Information Criterion.

**a.**

| Model | CI | LL null model | LL full model | AIC |
|---|---|---|---|---|
| **Demographics** | 0.6967168 | -66 860.64 | -65 346.33 | 130 698.7 |
| **+ RetFit** | 0.6982227 | | -65 320.64 | 130 655.3 |
| **+ SETCRF** | 0.6974730 | | -65 333.88 | 130 681.8 |
| **+ RetFit + SETCRF** | 0.6988471 | | -65 310.20 | 130 642.4 |
| **Demographics + CV** | 0.7207364 | -57 211.09 | -55 503.21 | 111 028.4 |
| **+ RetFit** | 0.7209531 | | -55 496.45 | 111 022.9 |
| **+ SETCRF** | 0.7208208 | | -55 500.78 | 111 031.6 |
| **+ RetFit + SETCRF** | 0.7210327 | | -55 494.06 | 111 026.1 |

**b.**

| Model | CI | LL null model | LL full model | AIC |
|---|---|---|---|---|
| **Demographics** | 0.7182870 | -42 689.65 | -41 460.70 | 82 927.39 |
| **+ RetFit** | 0.7203598 | | -41 436.37 | 82 886.73 |
| **+ SETCRF** | 0.7215751 | | -41 411.33 | 82 836.65 |
| **+ RetFit + SETCRF** | 0.7231764 | | -41 390.06 | 82 802.13 |
| **Demographics + CV** | 0.7367081 | -36 356.86 | -35 087.58 | 70 197.17 |
| **+ RetFit** | 0.7377271 | | -35 077.01 | 70 184.03 |
| **+ SETCRF** | 0.7383930 | | -35 055.79 | 70 141.57 |
| **+ RetFit + SETCRF** | 0.7392693 | | -35 045.92 | 70 129.84 |

**Suppl. Table 9 | Group survival analysis in the Rotterdam Study.** Group size and event distribution for the external and independent validation in the Rotterdam Study cohort. The thresholds reported in the table were computed independently in the UKBB dataset. Only people without previous cardiovascular events and with follow-up information were included in the survival analysis (n=6 110, out of N=8 885 subjects). CV: cardiovascular; UKBB: UK Biobank.

| **Group** | **RetFit Thresholds** | **Numerosity** | **CV Events (12y)** |
|---|---|---|---|
| Group 1 (least fit) | [0; 26.07) | 938 | 201 |
| Group 2 | [26.07; 27.62) | 1046 | 192 |
| Group 3 | [27.62; 29.20) | 1175 | 174 |
| Group 4 | [29.20; 31.20) | 1329 | 159 |
| Group 5 (most fit) | [31.20; inf) | 1622 | 129 |

**Suppl. Table 10 | Covariates used during the GWAS and corresponding UK Biobank data fields.**

| Covariate | UKB Data Field |
|---|---|
| Age | 21022 |
| Sex | 22001 |
| BMI | 21001 |
| UK Biobank assessment centre | 54 |
| Spherical power (left) | 5085 |
| Spherical power (right) | 5084 |
| Cylindrical power (left) | 5086 |
| Cylindrical power (right) | 5087 |
| Genotype measurement batch | 22000 |
| Genetic principal components | 22009 |

**Suppl. Table 11 | Complete results of the gene enrichment analysis in the UK Biobank. a)** Table showing all significant genes found for RetFit after Bonferroni correction, sorted by ascending p-values. **b)** Same as **a)**, but for SETCRF.

**a.**

| Gene | Chromosome | p-value |
|---|---|---|
| KIAA1755 | 20 | 4.291012e-14 |
| BPI | 20 | 1.965206e-11 |
| CCDC141 | 2 | 1.344749e-10 |
| CMTM5 | 14 | 1.047288e-09 |
| IL25 | 14 | 1.049337e-09 |
| EFS | 14 | 2.390951e-09 |
| MYH6 | 14 | 2.628005e-09 |
| SLC22A17 | 14 | 4.379828e-08 |
| TGM2 | 20 | 5.657446e-08 |
| THRB | 3 | 5.998647e-08 |
| CTD-2128A3.3 | 14 | 7.835316e-08 |
| LRRC37A | 17 | 2.319681e-07 |
| ARL17B | 17 | 2.380198e-07 |
| NKX2-5 | 5 | 2.494037e-07 |
| RP11-798G7.6 | 17 | 3.716767e-07 |
| LRRC53 | 1 | 4.026549e-07 |
| CTD-2128A3.2 | 14 | 4.775857e-07 |
| SPPL2C | 17 | 5.186473e-07 |
| MAPT | 17 | 5.246828e-07 |
| KANSL1 | 17 | 5.416399e-07 |
| STH | 17 | 5.707930e-07 |
| CRHR1 | 17 | 6.081518e-07 |
| RP11-293E1.2 | 17 | 7.431690e-07 |
| RP11-293E1.1 | 17 | 7.549612e-07 |
| SCN10A | 3 | 8.938093e-07 |

**b.**

| Gene | Chromosome | p-value |
|---|---|---|
| ACP1 | 2 | 2.469802e-48 |
| SH3YL1 | 2 | 6.760335e-47 |
| FAM150B | 2 | 5.882390e-45 |
| AC079779.5 | 2 | 8.074833e-43 |
| AC079779.6 | 2 | 1.221714e-38 |
| MTHFD1L | 6 | 4.184031e-23 |
| NPLOC4 | 17 | 8.154401e-19 |
| PDE6G | 17 | 3.955171e-18 |
| OXLD1 | 17 | 2.363888e-16 |
| CCDC137 | 17 | 2.509307e-16 |
| BCMO1 | 16 | 9.214851e-15 |
| ARL 16 | 17 | 5.073719e-14 |
| HGS | 17 | 1.704747e-13 |
| C17orf70 | 17 | 6.394885e-13 |
| FSCN2 | 17 | 4.862499e-12 |
| RLBP1 | 15 | 1.759393e-11 |
| ABHD2 | 15 | 3.622569e-11 |
| ACTG1 | 17 | 5.633818e-10 |
| AC139099.4 | 17 | 1.166346e-08 |
| DNAJC24 | 11 | 1.687606e-08 |
| ALDH6A1 | 14 | 2.021089e-08 |
| LIN52 | 14 | 2.129462e-08 |
| Z83307.3 | 11 | 3.751249e-08 |
| CCDC176 | 14 | 3.841253e-08 |
| FOXP1 | 3 | 6.721635e-08 |
| DCDC1 | 11 | 8.976101e-08 |
| SOX7 | 8 | 9.211016e-08 |

| | | |
|---|---|---|
| PINX1 | 8 | 9.348683e-08 |
| FANCI | 15 | 1.869548e-07 |
| PAX6 | 11 | 2.134046e-07 |
| AC144831.1 | 17 | 2.286861e-07 |
| RP11-1055B8.7 | 17 | 3.370605e-07 |
| DEPDC5 | 22 | 3.560447e-07 |
| RP11-554A11.8 | 11 | 4.652888e-07 |
| HMHA1 | 19 | 1.009941e-06 |
| RBBP5 | 1 | 1.093118e-06 |
| FAM110C | 2 | 1.559689e-06 |
| POLR2E | 19 | 1.700506e-06 |
| IMMP1L | 11 | 1.749795e-06 |
| CATSPER4 | 1 | 1.843093e-06 |
| PRR14L | 22 | 1.948590e-06 |

**Suppl. Table 12 | Complete results of the pathway analysis in the UK Biobank. a)** Table showing the top 10 significant pathways found for RetFit and their corresponding description extracted from https://www.gsea-msigdb.org/. **b)** Same as **a)**, but for SETCRF. **c)** Full table of all the significant pathways found for RetFit after Benjamini-Hochberg correction. **d)** Same as **c)**, but for SETCRF.

**a.**

| Pathway name | Number of genes involved | p-value | Description |
|---|---|---|---|
| HP_ABNORMALITY_OF_VISION | 827 | 2.5563581220392893e-07 | Abnormality of eyesight (visual perception) |
| LBP1_Q6 | 167 | 4.5082015865937693e-07 | Genes having at least one occurrence of the motif CAGCTGS in the regions spanning 4 kb centered on their transcription starting sites (-2kb to +2kb). This matches the UBP1 transcription factor binding site. |
| HP_ABNORMAL_CONJUGATE_EYE_MOVEMENT | 645 | 3.73211325333164e-06 | Any deviation from the normal motor coordination of the eyes that allows for bilateral fixation on a single object. |
| HP_CYSTOID_MACULAR_EDEMA | 15 | 5.251139081461455e-06 | Cystoid macular edema (CME) is any type of macular edema that involves cyst formation. |
| GO_REGULATION_OF_NEURON_DIFFERENTIATION | 550 | 5.478615715029208e-06 | Any process that modulates the frequency, rate, or extent of neuron differentiation. |
| GO_REGULATION_OF_CELL_MORPHOGENESIS_INVOLVED_IN_DIFFERENTIATION | 265 | 5.637606824006983e-06 | The change in form (cell shape and size) that occurs when relatively unspecialized cells, e.g., embryonic or regenerative cells, acquire specialized structural and/or functional features that characterize the cells, tissues, or organs of the mature organism or some other relatively stable phase of the organism's life history. |
| GO_REGULATION_OF_CELL_DEVELOPMENT | 781 | 7.1567271204386535e-06 | Any process that modulates the rate, frequency, or extent of the progression of the cell over time, from its formation to the mature structure. Cell development does not include the steps involved in committing a cell to a specific fate. |
| HP_ABNORMALITY_OF_THE_OPTIC_DISC | 464 | 1.2069735129154093e-05 | Abnormality of the optic disc. |
| ZFHX3_TARGET_GENES | 972 | 1.24573119368828e-05 | Genes containing one or more binding sites for UniProt:Q15911 (ZFHX3) in their promoter regions (TSS -1000,+100 bp) |
| ZNF596_TARGET_GENES | 439 | 1.2977732153016633e-05 | Genes containing one or more binding sites for UniProt:Q8TC21 (ZNF596) in their promoter regions (TSS -1000,+100 bp) |

**b.**

| Pathway name | Number of genes involved | p-value | Description |
|---|---|---|---|
| HP_CARDIAC_CONDUCTION_ABNORMALITY | 84 | 7.552741145306443e-12 | Any anomaly of the progression of electrical impulses through the heart. |
| HP_HEART_BLOCK | 74 | 2.502993246489469e-11 | Impaired conduction of cardiac impulse occurring anywhere along the conduction pathway. |
| GO_HEART_PROCESS | 228 | 4.480861491645533e-11 | A circulatory system process carried out by the heart. The heart is a hollow, muscular organ, which, by contracting rhythmically, keeps up the circulation of the blood. The heart is a hollow, muscular organ, which, by contracting rhythmically, keeps up the circulation of the blood. |
| HP_SUPRAVENTRICULAR_ARRHYTHMIA | 84 | 2.0598383175411944e-10 | A type of arrhythmia that originates above the ventricles, whereby the electrical impulse propagates down the normal His Purkinje system similar to normal sinus rhythm. |
| HP_ATRIAL_ARRHYTHMIA | 61 | 5.178328385942716e-10 | A type of supraventricular tachycardia in which the atria are the principal site of electrophysiologic disturbance. |
| GO_CARDIAC_MUSCLE_CONTRACTION | 111 | 1.184355235112929e-09 | Muscle contraction of cardiac muscle tissue. |
| GO_REGULATION_OF_STRIATED_MUSCLE_CONTRACTION | 73 | 1.2471449496900534e-09 | Any process that modulates the frequency, rate or extent of striated muscle contraction. |
| GO_REGULATION_OF_BLOOD_CIRCULATION | 231 | 1.4645266552922262e-09 | Any process that modulates the frequency, rate or extent of blood circulation. |
| GO_CIRCULATORY_SYSTEM_PROCESS | 450 | 2.0842756118687353e-09 | An organ system process carried out by any of the organs or tissues of the circulatory system. The circulatory system is an organ system that moves extracellular fluids to and from tissue within a multicellular organism. |
| GO_STRIATED_MUSCLE_CONTRACTION | 141 | 4.326893259731141e-09 | A process in which force is generated within striated muscle tissue, resulting in the shortening of the muscle. Force generation involves a chemo-mechanical energy conversion step that is carried out by the actin/myosin complex activity, which generates force through ATP hydrolysis. Striated muscle is a type of muscle in which the repeating units (sarcomeres) of the contractile myofibrils are arranged in registry throughout the cell, resulting in transverse or oblique striations observable at the level of the light microscope. |

**c.**

| Pathway name | Number of genes | p value |
|---|---|---|
| HP_ABNORMALITY_OF_VISION | 827 | 2.5563581220392893e-07 |
| LBP1_Q6 | 167 | 4.5082015865937693e-07 |
| HP_ABNORMAL_CONJUGATE_EYE_MOVEMENT | 645 | 3.73211325333164e-06 |
| HP_CYSTOID_MACULAR_EDEMA | 15 | 5.251139081461455e-06 |
| GO_REGULATION_OF_NEURON_DIFFERENTIATION | 550 | 5.478615715029208e-06 |
| GO_REGULATION_OF_CELL_MORPHOGENESIS_INVOLVED_IN_DIFFERENTIATION | 265 | 5.637606824006983e-06 |
| GO_REGULATION_OF_CELL_DEVELOPMENT | 781 | 7.1567271204386535e-06 |
| HP_ABNORMALITY_OF_THE_OPTIC_DISC | 464 | 1.2069735129154093e-05 |
| ZFHX3_TARGET_GENES | 972 | 1.24573119368828e-05 |
| ZNF596_TARGET_GENES | 439 | 1.2977732153016633e-05 |
| GSE17721_CTRL_VS_POLYIC_4H_BMDC_UP | 172 | 1.4157475476483524e-05 |
| ZNF843_TARGET_GENES | 548 | 1.4791086653758795e-05 |
| GGGYGTGNY_UNKNOWN | 515 | 1.582724650750508e-05 |
| HP_ABNORMAL_INVOLUNTARY_EYE_MOVEMENTS | 689 | 1.7894613983392477e-05 |
| HP_ABNORMAL_REPRODUCTIVE_SYSTEM_MORPHOLOGY | 859 | 1.8150840376439855e-05 |
| PXR_Q2 | 220 | 2.0620218947560463e-05 |
| HP_MACULAR_THICKENING | 22 | 2.2259682445519625e-05 |
| HP_ABNORMAL_EXTERNAL_GENITALIA | 717 | 2.3743393061171948e-05 |
| HP_ABNORMAL_FACIAL_EXPRESSION | 107 | 2.38073744032559e-05 |
| GSE15324_NAIVE_VS_ACTIVATED_ELF4_KO_CD8_TCELL_UP | 169 | 2.8645799593884432e-05 |
| ZNF592_TARGET_GENES | 1 045 | 3.0171887270827542e-05 |
| HP_ABNORMALITY_OF_THE_OPTIC_NERVE | 545 | 3.0594980338960915e-05 |
| GSE11924_TFH_VS_TH2_CD4_TCELL_UP | 164 | 3.539200725463967e-05 |
| HP_ABNORMAL_HAIR_PATTERN | 177 | 3.7294956106466696e-05 |
| HP_ABNORMALITY_OF_THE_CEREBRAL_SUBCORTEX | 662 | 3.845944293165199e-05 |
| HP_CONGENITAL_ABNORMAL_HAIR_PATTERN | 157 | 3.850466626949812e-05 |
| HP_ATTENUATION_OF_RETINAL_BLOOD_VESSELS | 37 | 4.098839224173155e-05 |
| GO_NEGATIVE_REGULATION_OF_NERVOUS_SYSTEM_DEVELOPMENT | 267 | 4.106604872864529e-05 |
| HP_ABNORMAL_FOVEAL_MORPHOLOGY | 35 | 4.284246466916859e-05 |

| | | |
|---|---|---|
| KYNG_ENVIRONMENTAL_STRESS_RESPONSE_NOT_BY_UV_IN_OLD | 23 | 4.872231346072348e-05 |
| KYNG_ENVIRONMENTAL_STRESS_RESPONSE_NOT_BY_GAMMA_IN_OLD | 29 | 5.0776672659133976e-05 |
| GO_REGULATION_OF_NERVOUS_SYSTEM_DEVELOPMENT | 762 | 5.149150403713333e-05 |
| SMN1_SMN2_TARGET_GENES | 528 | 5.24410253058506e-05 |
| GSE24210_TCONV_VS_TREG_DN | 165 | 5.403003560834547e-05 |
| GO_REGULATION_OF_CELL_MORPHOGENESIS | 417 | 5.46997793288935e-05 |
| GO_NEGATIVE_REGULATION_OF_NEURON_DIFFERENTIATION | 189 | 5.654258753370123e-05 |
| CBX7_TARGET_GENES | 602 | 5.988457621644188e-05 |
| HP_REDUCED_VISUAL_ACUITY | 349 | 6.14661702284534e-05 |
| HP_ABNORMAL_CORPUS_CALLOSUM_MORPHOLOGY | 522 | 6.262568820500083e-05 |

**d.**

| Pathway name | Number of genes | p value |
|---|---|---|
| HP_CARDIAC_CONDUCTION_ABNORMALITY | 84 | 7.552741145306443e-12 |
| HP_HEART_BLOCK | 74 | 2.502993246489469e-11 |
| GO_HEART_PROCESS | 228 | 4.480861491645533e-11 |
| HP_SUPRAVENTRICULAR_ARRHYTHMIA | 84 | 2.0598383175411944e-10 |
| HP_ATRIAL_ARRHYTHMIA | 61 | 5.178328385942716e-10 |
| GO_CARDIAC_MUSCLE_CONTRACTION | 111 | 1.184355235112929e-09 |
| GO_REGULATION_OF_STRIATED_MUSCLE_CONTRACTION | 73 | 1.2471449496900534e-09 |
| GO_REGULATION_OF_BLOOD_CIRCULATION | 231 | 1.4645266552922262e-09 |
| GO_CIRCULATORY_SYSTEM_PROCESS | 450 | 2.0842756118687353e-09 |
| GO_STRIATED_MUSCLE_CONTRACTION | 141 | 4.326893259731141e-09 |
| GO_REGULATION_OF_CARDIAC_MUSCLE_CONTRACTION | 63 | 4.8861057502471505e-09 |
| HP_ABNORMAL_LEFT_VENTRICLE_MORPHOLOGY | 77 | 1.1872341337961656e-08 |
| GO_MUSCLE_SYSTEM_PROCESS | 357 | 1.5429306154122234e-08 |
| MODULE_201 | 48 | 2.60825768092578e-08 |
| HP_BUNDLE_BRANCH_BLOCK | 41 | 2.701346793987184e-08 |
| HP_ABNORMALITY_OF_CARDIOVASCULAR_SYSTEM_ELECTROPHYSIOLOGY | 329 | 2.9511659254251763e-08 |
| HP_ABNORMAL_CARDIOVASCULAR_SYSTEM_PHYSIOLOGY | 827 | 3.1608663447839273e-08 |
| BRUNEAU_SEPTATION_ATRIAL | 5 | 5.46410150849701e-08 |
| GO_CARDIAC_CONDUCTION | 114 | 5.637188247332607e-08 |
| GO_MUSCLE_CONTRACTION | 287 | 6.019984318242944e-08 |
| REACTOME_MUSCLE_CONTRACTION | 162 | 9.245965457335369e-08 |
| GO_REGULATION_OF_SYSTEM_PROCESS | 480 | 1.0420089666344285e-07 |
| HP_LEFT_VENTRICULAR_HYPERTROPHY | 59 | 1.0969557666049345e-07 |
| HP_VENTRICULAR_HYPERTROPHY | 88 | 1.9755734997940315e-07 |
| GO_REGULATION_OF_HEART_RATE | 83 | 2.2530869933697018e-07 |
| GO_MULTICELLULAR_ORGANISMAL_SIGNALING | 164 | 2.389474563317444e-07 |
| GO_REGULATION_OF_MUSCLE_SYSTEM_PROCESS | 187 | 3.328140154407002e-07 |
| HP_CONGESTIVE_HEART_FAILURE | 152 | 4.403401426053197e-07 |

| | | |
|---|---|---|
| GO_CELL_COMMUNICATION_INVOLVED_IN_CARDIAC_CONDUCTION | 47 | 4.66283746388333e-07 |
| HP_BRADYCARDIA | 50 | 6.703671552169626e-07 |
| HP_ABNORMAL_ATRIOVENTRICULAR_CONDUCTION | 53 | 6.853903899842009e-07 |
| REACTOME_CARDIAC_CONDUCTION | 110 | 7.743261709757143e-07 |
| HP_INCREASED_BLOOD_PRESSURE | 276 | 9.453921279962782e-07 |
| MODULE_329 | 48 | 1.15830384922002e-06 |
| GO_MUSCLE_CELL_DEVELOPMENT | 148 | 1.3929133241110135e-06 |
| HP_LEFT_ANTERIOR_FASCICULAR_BLOCK | 5 | 1.594842516859426e-06 |
| HP_SUDDEN_DEATH | 21 | 1.633418791303168e-06 |
| HP_VENTRICULAR_ARRHYTHMIA | 76 | 1.719330447688541e-06 |
| HP_TACHYCARDIA | 84 | 1.7909009313622262e-06 |
| MODULE_387 | 46 | 2.2874481371347595e-06 |
| HP_LEFT_BUNDLE_BRANCH_BLOCK | 8 | 2.309241665605702e-06 |
| GO_MUSCLE_ADAPTATION | 91 | 2.5087385836550988e-06 |
| HP_ABNORMAL_CARDIAC_VENTRICLE_MORPHOLOGY | 388 | 2.919079318427111e-06 |
| GO_MUSCLE_FIBER_DEVELOPMENT | 51 | 3.1839784925395293e-06 |
| HP_VENTRICULAR_TACHYCARDIA | 46 | 3.7812158037281406e-06 |
| GO_CARDIAC_MUSCLE_CELL_CONTRACTION | 60 | 3.7961315295497615e-06 |
| GO_REGULATION_OF_ACTIN_FILAMENT_BASED_MOVEMENT | 36 | 4.027800238891605e-06 |
| GO_REGULATION_OF_MUSCLE_CONTRACTION | 133 | 4.389855138108844e-06 |
| LAKE_ADULT_KIDNEY_C13_THICK_ASCENDING_LIMB | 110 | 5.367453574853714e-06 |
| HP_VENTRICULAR_EXTRASYSTOLES | 10 | 5.905140153438569e-06 |
| HP_ABNORMAL_SYSTEMIC_BLOOD_PRESSURE | 331 | 7.094594457801495e-06 |
| HP_RIGHT_BUNDLE_BRANCH_BLOCK | 34 | 8.213117700205589e-06 |
| HP_FIRST_DEGREE_ATRIOVENTRICULAR_BLOCK | 23 | 8.341379593699105e-06 |
| GO_CALMODULIN_BINDING | 173 | 8.4487797297238e-06 |
| GO_ORGAN_GROWTH | 151 | 8.956256640191881e-06 |
| GO_REGULATION_OF_MUSCLE_ADAPTATION | 71 | 9.781470900201286e-06 |
| HP_ABNORMALITY_OF_THE_MUSCULATURE_OF_THE_HAND | 57 | 1.002233646128272e-05 |
| GO_CARDIAC_MUSCLE_FIBER_DEVELOPMENT | 9 | 1.0233414563134648e-05 |
| chr17q21 | 7 | 1.3068154924792033e-05 |

| | | |
|---|---|---|
| PID_AVB3_OPN_PATHWAY | 29 | 1.3148341218837536e-05 |
| GO_REGULATION_OF_HEART_GROWTH | 62 | 1.3197648454773094e-05 |
| HP_PROLONGED_QTC_INTERVAL | 24 | 1.3458691771495978e-05 |
| HP_SINUS_BRADYCARDIA | 19 | 1.410819358513136e-05 |
| GO_CELL_CELL_SIGNALING_INVOLVED_IN_CARDIAC_CONDUCTION | 25 | 1.433147637254276e-05 |
| MODULE_1 | 309 | 1.4690701317248912e-05 |
| GO_ACTIN_MEDIATED_CELL_CONTRACTION | 95 | 1.6871312885825984e-05 |
| GO_CARDIAC_MUSCLE_CELL_ACTION_POTENTIAL | 58 | 1.8349191843104225e-05 |
| HALLMARK_HYPOXIA | 165 | 2.1486533137538142e-05 |
| GO_AV_NODE_CELL_TO_BUNDLE_OF_HIS_CELL_COMMUNICATION | 9 | 2.2315238569500447e-05 |
| GO_MYOFIBRIL_ASSEMBLY | 60 | 2.3720510389850436e-05 |
| GO_SARCOMERE_ORGANIZATION | 40 | 2.427141031479154e-05 |
| GO_HEART_GROWTH | 83 | 2.467563159723265e-05 |
| CUI_TCF21_TARGETS_DN | 27 | 2.520520540025379e-05 |
| HP_TIBIALIS_MUSCLE_WEAKNESS | 5 | 2.623584674061441e-05 |
| GO_REGULATION_OF_HEART_RATE_BY_CARDIAC_CONDUCTION | 34 | 2.8810675304965197e-05 |
| GO_REGULATION_OF_THE_FORCE_OF_HEART_CONTRACTION | 22 | 2.909319955787365e-05 |
| LIN_APC_TARGETS | 59 | 3.0842735600875426e-05 |
| HP_ABNORMAL_ATRIOVENTRICULAR_VALVE_MORPHOLOGY | 89 | 3.285608256394013e-05 |
| CACCAGC_MIR138 | 187 | 3.3404618191501706e-05 |
| GO_PLASMA_MEMBRANE_PROTEIN_COMPLEX | 427 | 3.3796004056633874e-05 |
| REACTOME_FOXO_MEDIATED_TRANSCRIPTION | 60 | 3.412340312369083e-05 |
| GO_MEMBRANE_DEPOLARIZATION_DURING_AV_NODE_CELL_ACTION_POTENTIAL | 5 | 3.463980988241218e-05 |
| GO_BUNDLE_OF_HIS_CELL_TO_PURKINJE_MYOCYTE_COMMUNICATION | 13 | 3.480018499482964e-05 |
| HP_SUPRAVENTRICULAR_TACHYCARDIA | 30 | 3.589100569748411e-05 |
| GO_ENDOCRINE_PROCESS | 69 | 3.667211502647189e-05 |
| TCCATTKW_UNKNOWN | 196 | 3.678016334086895e-05 |
| HP_ABNORMAL_CARDIAC_TEST | 82 | 3.8382149786640495e-05 |
| GO_DEVELOPMENTAL_GROWTH | 538 | 4.191148330612305e-05 |
| PID_LYSOPHOSPHOLIPID_PATHWAY | 59 | 4.225498223696373e-05 |
| GO_MUSCLE_HYPERTROPHY | 76 | 4.331470204758361e-05 |

| | | |
|---|---|---|
| HP_SUBVALVULAR_AORTIC_STENOSIS | 14 | 4.418585782279781e-05 |
| GO_REGULATION_OF_ORGAN_GROWTH | 88 | 4.463583180773454e-05 |
| HP_ABNORMAL_MITRAL_VALVE_MORPHOLOGY | 72 | 4.807093238850101e-05 |
| GO_ACTION_POTENTIAL | 112 | 4.856004012978658e-05 |
| HP_MANDIBULAR_PROGNATHIA | 121 | 4.9262859865776485e-05 |
| GO_RESPONSE_TO_OXYGEN_LEVELS | 326 | 5.279548946869818e-05 |
| GO_ACTIN_FILAMENT_BASED_MOVEMENT | 123 | 5.393626668682679e-05 |
| GO_CHROMATIN_BINDING | 460 | 5.665805763844866e-05 |
| MIR6865_3P | 38 | 6.1251268773116e-05 |
| GO_ADULT_HEART_DEVELOPMENT | 12 | 6.372039803869511e-05 |
| GO_CARDIAC_MUSCLE_TISSUE_DEVELOPMENT | 189 | 6.620929851756702e-05 |
| TGACATY_UNKNOWN | 537 | 7.014105671145237e-05 |
| MIR4261 | 355 | 7.17576522813776e-05 |
| GO_BEHAVIOR | 497 | 7.242517370408074e-05 |
| GO_MUSCLE_TISSUE_DEVELOPMENT | 334 | 7.707286838285579e-05 |
| GO_REGULATION_OF_CARDIAC_CONDUCTION | 54 | 8.757762672953863e-05 |
| GSE15930_NAIVE_VS_24H_IN_VITRO_STIM_CD8_TCELL_UP | 173 | 8.819957816002723e-05 |
| HP_SECUNDUM_ATRIAL_SEPTAL_DEFECT | 9 | 9.170193328449234e-05 |
| CUI_DEVELOPING_HEART_C2_CARDIOMYOCYTE | 77 | 0.0001046026247404 |
| HP_PATCHY_PALMOPLANTAR_KERATODERMA | 3 | 0.0001046417714665 |
| HP_PROXIMAL_MUSCLE_WEAKNESS_IN_UPPER_LIMBS | 32 | 0.0001084337569936 |
| GO_CARDIAC_CELL_DEVELOPMENT | 69 | 0.0001102546976633 |
| GATA1_02 | 206 | 0.0001114072316319 |
| GO_GROWTH | 753 | 0.0001185483616552 |
| HP_ABNORMALITY_OF_CHROMOSOME_SEGREGATION | 11 | 0.0001201598113253 |
| GSE46606_IRF4HIGH_VS_WT_CD40L_IL2_IL5_DAY3_STIMULATED_BCELL_UP | 169 | 0.0001217165421862 |
| GO_ACTIN_FILAMENT_BASED_PROCESS | 630 | 0.0001236555186247 |
| GO_NEGATIVE_REGULATION_OF_MULTICELLULAR_ORGANISMAL_PROCESS | 942 | 0.0001255729591619 |
| HP_PALMOPLANTAR_KERATODERMA | 121 | 0.0001264666054924 |
| GO_FEMALE_MATING_BEHAVIOR | 8 | 0.0001269682352486 |
| CUI_DEVELOPING_HEART_CARDIAC_FIBROBLASTS | 88 | 0.0001274923873429 |

| | | |
|---|---|---|
| HP_GENERALIZED_HYPOTONIA | 691 | 0.0001295760229729 |
| WP_MYOMETRIAL_RELAXATION_AND_CONTRACTION_PATHWAYS | 137 | 0.0001342097753651 |
| GO_KINASE_BINDING | 618 | 0.0001375221821285 |
| REACTOME_PHYSIOLOGICAL_FACTORS | 11 | 0.0001456904933051 |
| HP_LEUKODYSTROPHY | 86 | 0.0001489288871517 |
| HP_NEUROFIBRILLARY_TANGLES | 20 | 0.0001508770315846 |
| HP_ABNORMALITY_OF_THE_BASAL_GANGLIA | 146 | 0.0001516461109146 |
| GO_CELLULAR_COMPONENT_ASSEMBLY_INVOLVED_IN_MORPHOGENESIS | 98 | 0.0001554454196056 |
| BRUNEAU_SEPTATION_VENTRICULAR | 10 | 0.0001573068365789 |
| HP_MUSCLE_FIBER_SPLITTING | 13 | 0.0001629234932974 |
| HP_TRICEPS_WEAKNESS | 15 | 0.0001642574549718 |
| chr6q22 | 21 | 0.00016537987501 |
| GO_ATRIAL_CARDIAC_MUSCLE_CELL_TO_AV_NODE_CELL_SIGNALING | 15 | 0.0001657704338133 |
| HP_HETEROGENEOUS | 220 | 0.0001684100195537 |
| GO_REGULATION_OF_MEMBRANE_POTENTIAL | 341 | 0.0001756336223703 |
| MIR518D_5P_MIR518F_5P_MIR520C_5P_MIR526A_5P | 81 | 0.0001760227143524 |
| KEGG_VASCULAR_SMOOTH_MUSCLE_CONTRACTION | 97 | 0.000178984478542 |
| GO_CARDIAC_MUSCLE_CELL_ACTION_POTENTIAL_INVOLVED_IN_CONTRACTION | 44 | 0.0001818643257854 |
| MIR6728_5P | 60 | 0.0001830552799596 |
| HP_ABNORMALITY_OF_THE_MUSCULATURE_OF_THE_UPPER_LIMBS | 140 | 0.000187855812577 |
| STAT5A_01 | 205 | 0.0001898318856432 |
| GO_REGULATION_OF_CARDIAC_MUSCLE_CELL_ACTION_POTENTIAL | 23 | 0.0001916226531906 |
| HP_ABNORMAL_LEFT_VENTRICULAR_OUTFLOW_TRACT_MORPHOLOGY | 17 | 0.0001953541305283 |
| HP_DISTAL_UPPER_LIMB_MUSCLE_WEAKNESS | 12 | 0.0001959577037138 |
| WP_CORTICOTROPINRELEASING_HORMONE_SIGNALING_PATHWAY | 85 | 0.0002011725797061 |
| MODULE_330 | 26 | 0.0002045028703593 |
| HP_PAIN | 474 | 0.000206438184104 |
| GO_THYROID_HORMONE_MEDIATED_SIGNALING_PATHWAY | 5 | 0.0002086151762426 |
| MIR147B_5P | 264 | 0.0002089349741982 |
| SIRNA_EIF4GI_UP | 82 | 0.0002144527201588 |
| GO_ACTOMYOSIN_STRUCTURE_ORGANIZATION | 173 | 0.0002153794859636 |

| | | |
|---|---|---|
| HP_DISTAL_LOWER_LIMB_MUSCLE_WEAKNESS | 45 | 0.0002180912226711 |
| CHIBA_RESPONSE_TO_TSA | 36 | 0.0002184746536089 |
| GNF2_MYL3 | 27 | 0.0002252683174662 |
| PID_AP1_PATHWAY | 65 | 0.0002256552658218 |
| GO_TISSUE_MORPHOGENESIS | 546 | 0.0002305022552635 |
| HP_DYSPAREUNIA | 33 | 0.0002307738290017 |

**Suppl. Table 13 | List and brief description of the Tangible Image Features (TIFs) used for the model interpretability analysis in the UK Biobank.**

| Feature Name | Abbreviation | Short Description |
|---|---|---|
| A temporal angle | ta_arteries | Angle of the temporal branch of the retinal artery |
| V temporal angle | ta_veins | Angle of the temporal branch of the retinal vein |
| A tortuosity | tort_arteries | Measure of twisting or curvature of retinal arteries |
| V tortuosity | tort_veins | Measure of twisting or curvature of retinal veins |
| Ratio tortuosity | tort_ratio | Ratio of arterial to venous tortuosity |
| A central retinal equivalent | cre_arteries | Average diameter of the central retinal arteries |
| V central retinal equivalent | cre_veins | Average diameter of the central retinal veins |
| Ratio central retinal equivalent | cre_ratio | Ratio of arterial to venous central retinal eq. diameters |
| A median diameter | diam_arteries_median | Median diameter of arteries |
| V median diameter | diam_veins_median | Median diameter of veins |
| Ratio median diameter | diam_ratio | Ratio of arterial to venous median diameter |
| A diameter std | diam_arteries_std | Measure of arterial diameter variability |
| V diameter std | diam_veins_std | Measure of venous diameter variability |
| A vascular density | vd_arteries | Proportion of the retinal area occupied by arteries |
| V vascular density | vd_veins | Proportion of the retinal area occupied by veins |
| Ratio vascular density | vd_ratio | Ratio of arterial to venous vascular density |
| A bifurcations | bif_arteries | Number of branch points in the retinal arteries |
| V bifurcations | bif_veins | Number of branch points in the retinal veins |

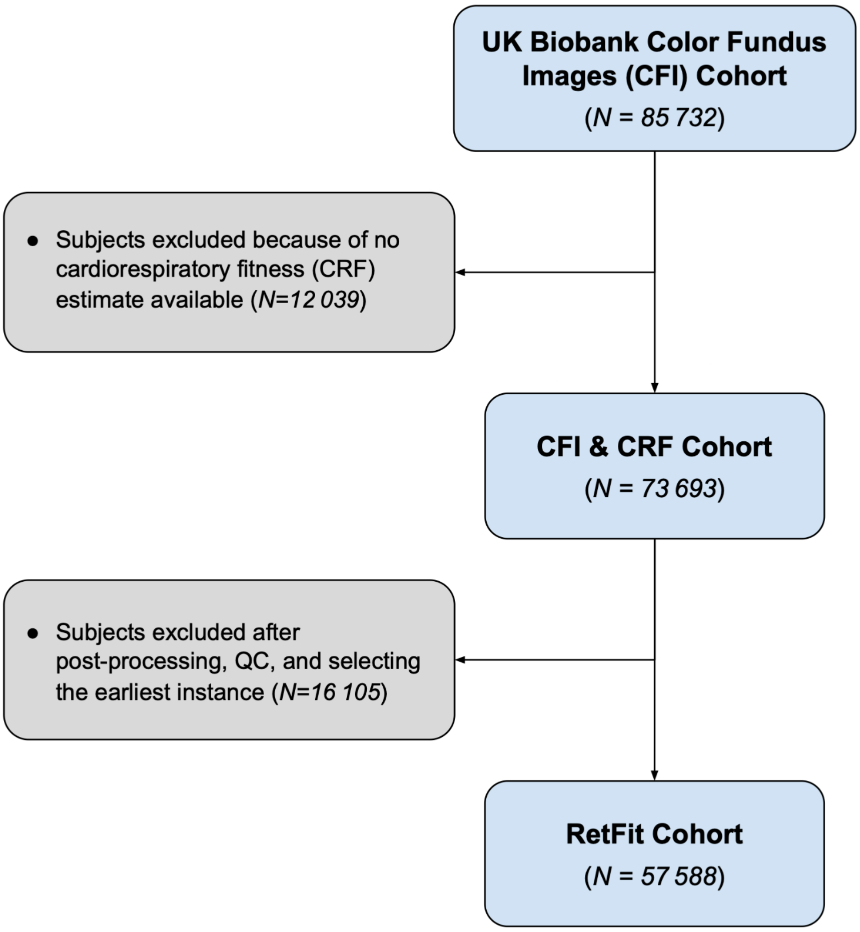


**Suppl. Figure 1 | CONSORT-style diagram for the UK Biobank analysis cohort.** We started from 85 732 subjects that got imaged, 73 693 of which had a cardiorespiratory fitness estimate. After quality control (see Methods in the manuscript), we were left with 56 876 images from instance 0 and 16 817 from instance 1. By selecting the earliest available instance for each subject (aiming for longer follow-up), and including every subject once, we were left with a final cohort of 57 588.

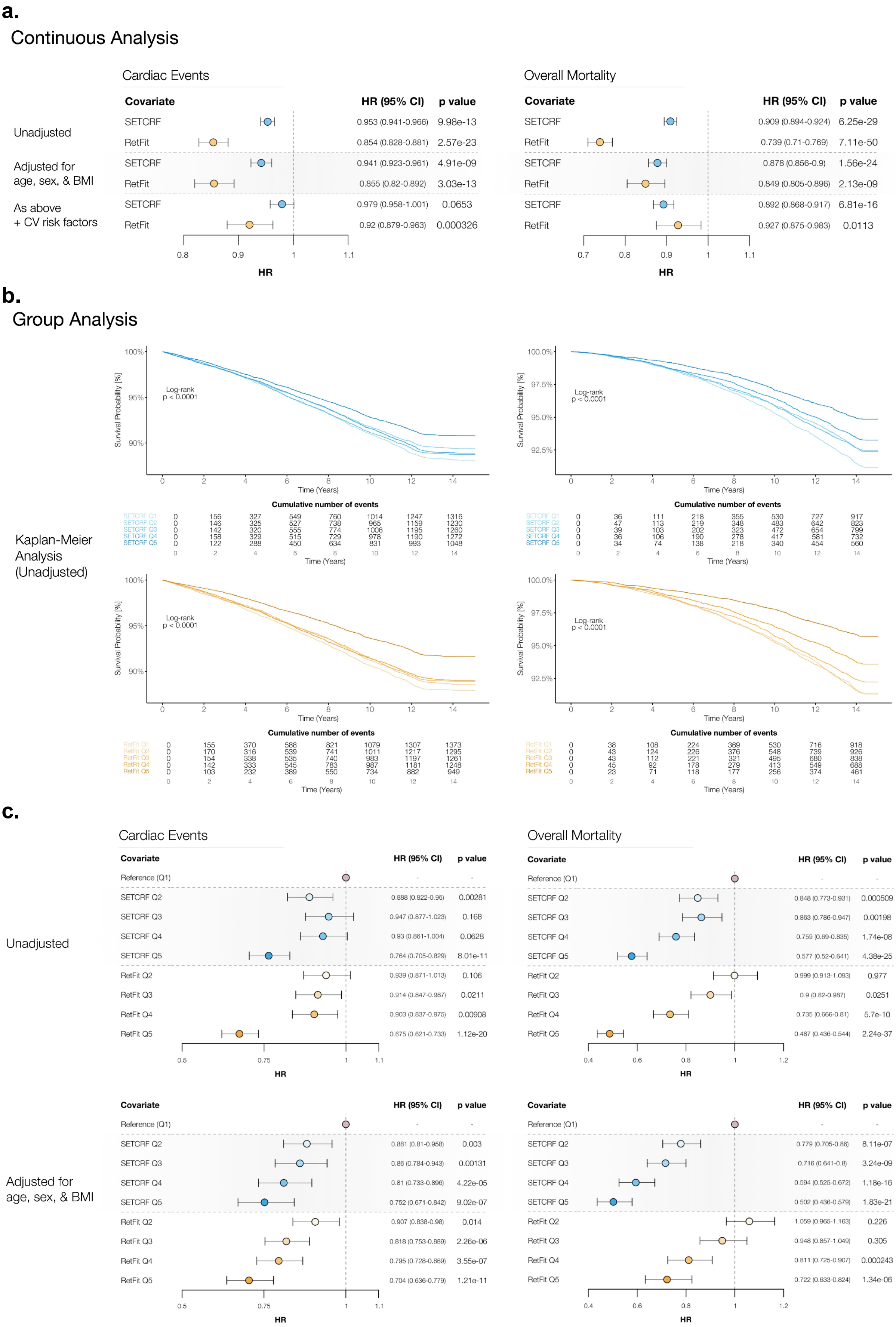

a.
Continuous Analysis
Cardiac Events
Covariate HR (95% CI) p value
Unadjusted
SETCRF 0.953 (0.941-0.966) 9.98e-13
RetFit 0.854 (0.828-0.881) 2.57e-23
Adjusted for age, sex, & BMI
SETCRF 0.941 (0.923-0.961) 4.91e-09
RetFit 0.855 (0.82-0.892) 3.03e-13
As above + CV risk factors
SETCRF 0.979 (0.958-1.001) 0.0653
RetFit 0.92 (0.879-0.963) 0.000326
HR
Overall Mortality
Covariate HR (95% CI) p value
SETCRF 0.909 (0.894-0.924) 6.25e-29
RetFit 0.739 (0.71-0.769) 7.11e-50
SETCRF 0.878 (0.856-0.9) 1.56e-24
RetFit 0.849 (0.805-0.896) 2.13e-09
SETCRF 0.892 (0.868-0.917) 6.81e-16
RetFit 0.927 (0.875-0.983) 0.0113
HR
b.
Group Analysis
Kaplan-Meier Analysis (Unadjusted)
Log-rank p < 0.0001
Survival Probability [%]
Time (Years)
Cumulative number of events
c.
Cardiac Events
Covariate HR (95% CI) p value
Unadjusted
Reference (Q1) - -
SETCRF Q2 0.888 (0.822-0.96) 0.00281
SETCRF Q3 0.947 (0.877-1.023) 0.168
SETCRF Q4 0.93 (0.861-1.004) 0.0628
SETCRF Q5 0.764 (0.705-0.829) 8.01e-11
RetFit Q2 0.939 (0.871-1.013) 0.106
RetFit Q3 0.914 (0.847-0.987) 0.0211
RetFit Q4 0.903 (0.837-0.975) 0.00908
RetFit Q5 0.675 (0.621-0.733) 1.12e-20
HR
Overall Mortality
Covariate HR (95% CI) p value
Reference (Q1) - -
SETCRF Q2 0.848 (0.773-0.931) 0.000509
SETCRF Q3 0.863 (0.786-0.947) 0.00198
SETCRF Q4 0.759 (0.69-0.835) 1.74e-08
SETCRF Q5 0.577 (0.52-0.641) 4.38e-25
RetFit Q2 0.999 (0.913-1.093) 0.977
RetFit Q3 0.9 (0.82-0.987) 0.0251
RetFit Q4 0.735 (0.666-0.81) 5.7e-10
RetFit Q5 0.487 (0.436-0.544) 2.24e-37
HR
Adjusted for age, sex, & BMI
Covariate HR (95% CI) p value
Reference (Q1) - -
SETCRF Q2 0.881 (0.81-0.958) 0.003
SETCRF Q3 0.86 (0.784-0.943) 0.00131
SETCRF Q4 0.81 (0.733-0.896) 4.22e-05
SETCRF Q5 0.752 (0.671-0.842) 9.02e-07
RetFit Q2 0.907 (0.838-0.98) 0.014
RetFit Q3 0.818 (0.753-0.889) 2.26e-06
RetFit Q4 0.795 (0.728-0.869) 3.55e-07
RetFit Q5 0.704 (0.636-0.779) 1.21e-11
HR
Covariate HR (95% CI) p value
Reference (Q1) - -
SETCRF Q2 0.779 (0.705-0.86) 8.11e-07
SETCRF Q3 0.716 (0.641-0.8) 3.24e-09
SETCRF Q4 0.594 (0.525-0.672) 1.18e-16
SETCRF Q5 0.502 (0.436-0.579) 1.83e-21
RetFit Q2 1.059 (0.965-1.163) 0.226
RetFit Q3 0.948 (0.857-1.049) 0.305
RetFit Q4 0.811 (0.725-0.907) 0.000243
RetFit Q5 0.722 (0.633-0.824) 1.34e-06
HR

**Suppl. Figure 2 | Prognostic performance of RetFit and SETCRF. a)** Analogous to **Fig. 3a**, forest plots of the CRF estimates as a continuous parameter for different clinical endpoints, but expressed in terms of METs. **b)** Kaplan-Meier survival analysis of the RetFit and SETCRF (all the quintiles, graphed separately for ease of visualization), and **c)** Forest plots of the RetFit and SETCRF quintiles, unadjusted and adjusted for demographics (age, sex, and BMI). HR: hazard ratio; CI: confidence interval; CV: cardiovascular.

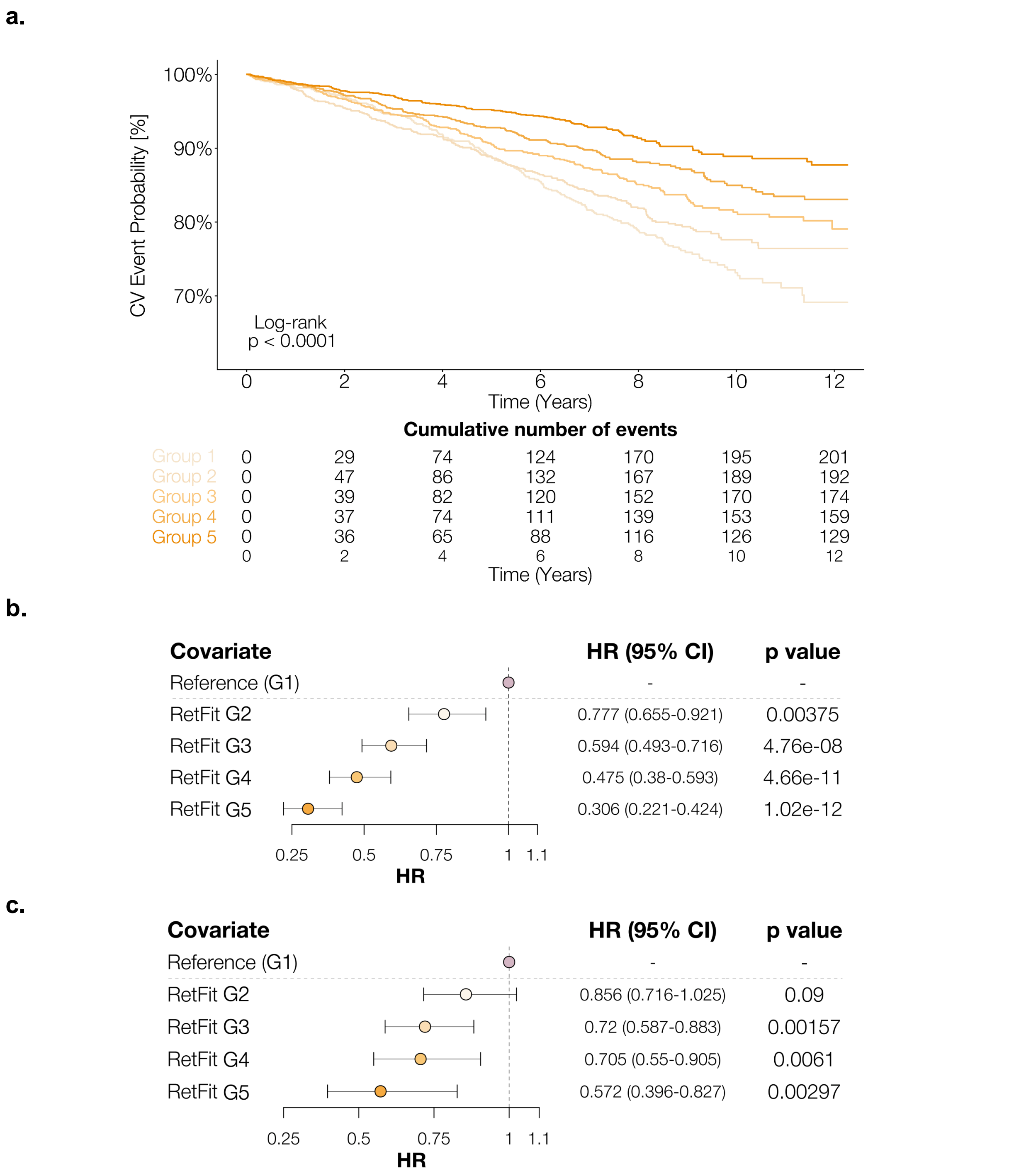


**Suppl. Figure 3 | Prognostic performance of RetFit in the Rotterdam Cohort. a)** Kaplan-Meier survival analysis of RetFit (all the groups). **b)** Forest plots of the RetFit groups, unadjusted, and **c)** adjusted for demographics (age, sex, and BMI). HR: hazard ratio; CI: confidence interval; CV: cardiovascular.

a.

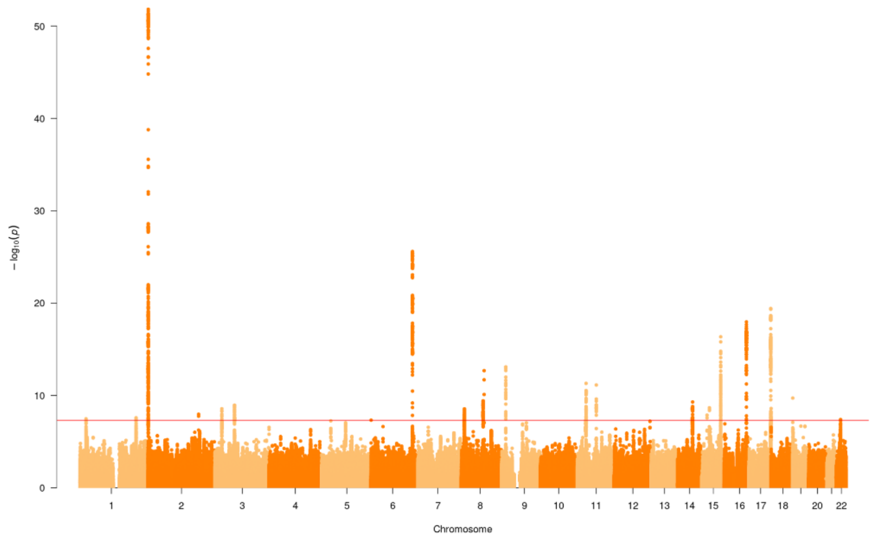


b.

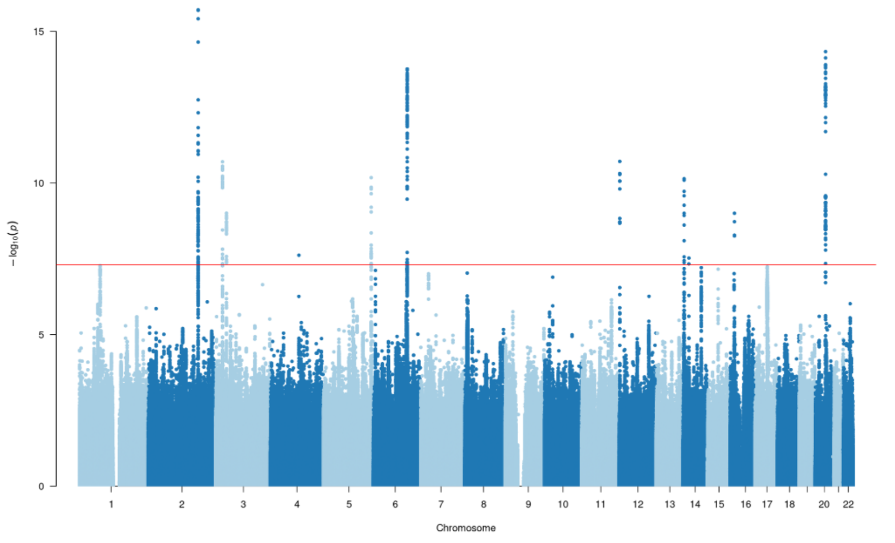


**Suppl. Figure 4 | Genome-wide SNPs Manhattan plots.** Manhattan plots obtained after running the Genome Wide Association Study for each of the two traits. Every dot on the graph represents a Single-Nucleotide Polymorphism (SNP). The red line represents the genome-wide significance set at 5e-8. **a)** SNP Manhattan plot for RetFit. **b)** SNP Manhattan plot for SETCRF.

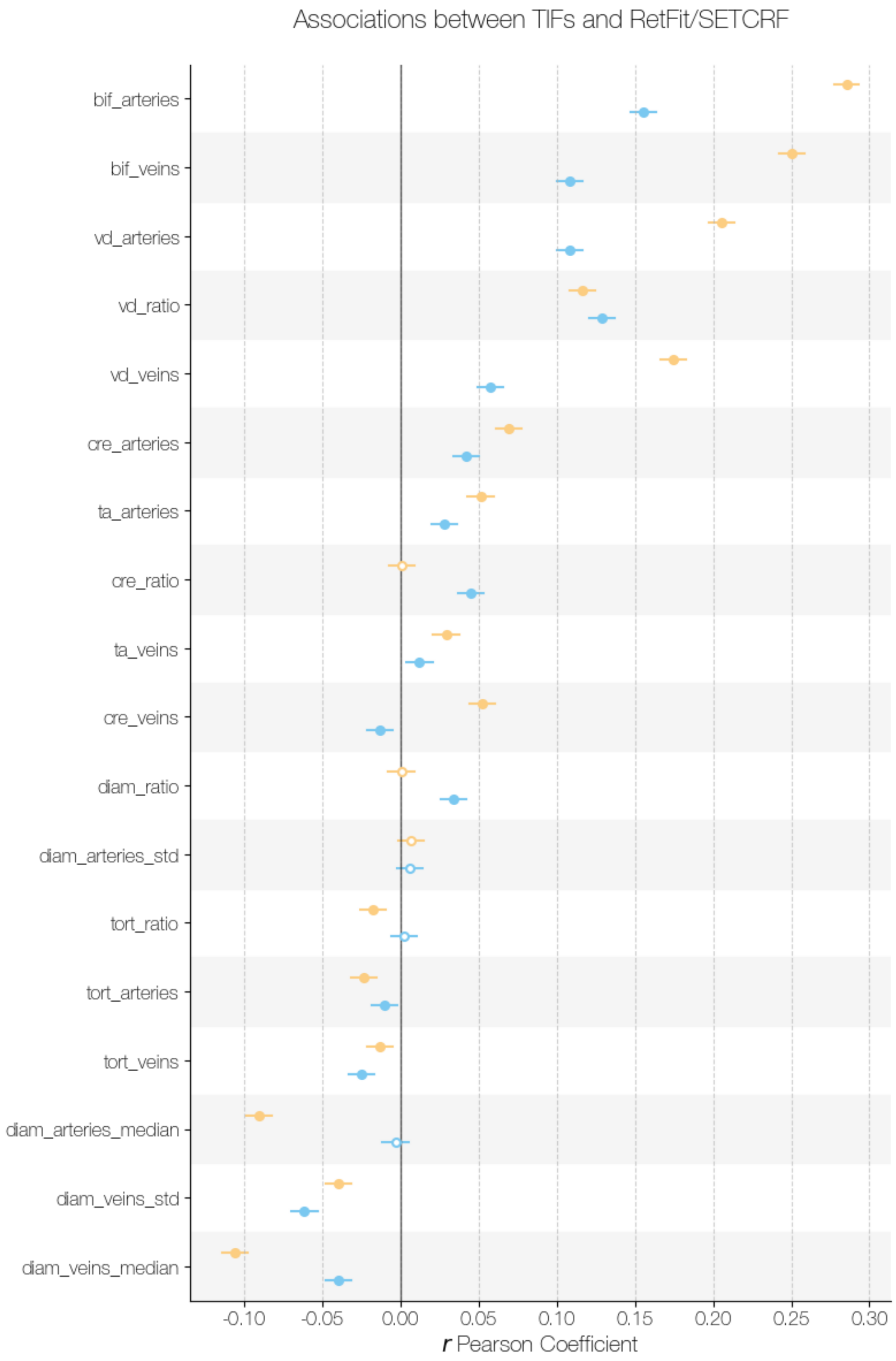


**Suppl. Figure 5 | Linear associations between RETFIT and SETCRF and retinal vascular features.** Forest plot showing Pearson correlation coefficients between features extracted from CFIs using VascX models, representing the vasculature of the retina, and RetFit and SETCRF.
